# ECHO: A lightweight tool for inferring missing case counts from pathogen phylogenies

**DOI:** 10.64898/2026.08.09.26360045

**Authors:** Renny Doig, Caroline Colijn

## Abstract

Timed phylogenetic trees express the evolutionary history of a pathogen outbreak in units of time, providing an estimate of the elapsed time across the shared ancestry of a set of taxa. By combining this elapsed time with known information about the epidemiology of a disease, we can relate the total branch length to the total number of cases related to the phylogeny. This gives information about the number of unsequenced cases that are related to the phylogeny. We call these “cryptic” cases. We present ECHO (Estimation of Cryptic Hosts from Outbreak trees), a collection of three lightweight estimators of the number of cryptic cases in a phylogeny. ECHO is agnostic to the form of the sampling process, making it robust to a variety of forms of sampling heterogeneity. We demonstrate ECHO’s baseline accuracy and its robustness to heterogenous sampling frameworks through simulation. Additionally, we apply ECHO to measles virus sequences that were collected during an outbreak in the USA in 2021. ECHO is able to recover the number of cryptic cases with a reasonable degree of accuracy both in simulation and in practice. We discuss the contexts in which ECHO is most applicable, and the interpretation of its estimates.

## Introduction

Genomic pathogen surveillance seldom leads to sampling and sequencing from all infected individuals, except perhaps in the most controlled settings. A good understanding of the sampling coverage is important for managing disease outbreaks. Despite this, relatively little of the epidemiology literature addresses estimation of the number of unsampled cases in an outbreak. Furthermore, as the accessibility of sequencing technology has increased, phylogenetic trees have become a standard tool for infectious disease surveillance and outbreak analysis in many public health jurisdictions. Consequently, using phylogenetic information for carrying out quantitative epidemiological analyses is potentially of broad interest to public health.

Phylogenetic reconstruction typically yields a tree whose branch lengths measure genetic divergence, expressed as the number of allele substitutions per site. Molecular clock models allow us to estimate the rate of mutation and create timed phylogenetic trees, whose branches are in units of calendar time. In the context of pathogen surveillance and genomics, this relationship allows the timed phylogeny to be linked to transmission events and transmission dynamics. This link can be leveraged to infer epidemiological quantities, including information about the sampling patterns and numbers of missing cases, from genomic data.

Relating the branching structure of the phylogeny to the spread of a pathogen in a population can be done using branching processes, in particular birth-death(-sampling) processes [1]. These models account for sampling, specifying the process by which it occurs [2, 3, 4, 5, 6]. While this can be used to account for a variety of types of sampling, not just uniform, fixed-rate sampling, it does require that the sampling process be known. These models are usually not robust to misspecification of this process. Another class of methods use the phylogenetic tree to reconstruct the transmission network of an outbreak [7, 8, 9]. These methods tend to require computationally intensive algorithms to infer the placement and timing of missing cases, rather than just their number. Furthermore, they are designed to reconstruct person-to-person transmission in densely sequenced outbreaks; most genomic surveillance efforts do not provide this level of resolution. However, in many infectious diseases, the distributions of the latent period (if there is one) and the infectious period are well established, and provide a natural connection between the number of intermediate transmissions and the edge lengths in timed phylogenies [10, 11].

In principle, given a timed phylogenetic tree and information about the natural history of infection, it should be possible to estimate the number of cases that are associated with that tree. To that end, we present ECHO (Estimation of Cryptic Hosts from Outbreak trees) as a lightweight tool for inferring the total number of unsampled infections related to a timed phylogenetic tree. The model is simple, relating the total branch length of the timed tree to known information about the progression of infection. This simplicity imparts a robustness to the sampling process: ECHO requires no prior information about the sampling fraction and is resilient to non-uniform sampling. We empirically demonstrate the accuracy and robustness of ECHO through several simulation experiments. A practical demonstration of ECHO is also performed using measles virus (MeV) sequences that were collected during an outbreak in the United States of America (USA) in 2021 [12].

## Methods

### ECHO

ECHO has three statistical models that relate the observed phylogeny and known epidemiology to the number of known and missing hosts related to the phylogeny. Suppose that *n*_obs_ sequences of a pathogen have been collected from different hosts and a timed phylogenetic tree *P* has been reconstructed from those sequences (we assume that exactly one sequence is observed per host). Because ECHO will take *P* as input, rather than inferring *P*, we refer to this as the “observed phylogeny”. *P* contains the topology of the phylogeny as well as the branch lengths; for the sake of this paper, we treat branch lengths as being measured in days. The total branch length is denoted |*P*|. The methods we derive in this paper rely on the common phylodynamic approximation that a branching event in *P* corresponds to an infection event in the population. In particular, say that there is a branching event corresponding to individual *P*_*j*_ infecting individual *j*; the clade associated with *j*’s lineage is denoted *P*_*j*_.

To denote the times at which events occur we use *t* for times referenced from the origin of the outbreak and *τ* for events with a different reference time. The progression of individual *i*’s infection is as follows: *i* is infected at time 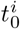, remains latent for 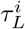 days, then is infectious for 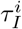 days. The infection lasts from 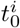 until 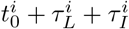. During the infectious period, *i* will infect new hosts; the set of all infectees of *i* are represented by *C*_*i*_. An infection event occurs at time 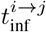 for *j* ∈ *C*_*i*_ and produces clade *P*_*j*_.

### Cryptic hosts

Whenever the sampling fraction is less than one, there will be some number of cases that are epidemiologically connected to an observed outbreak that are missing. Here we consider a case missing if it is not represented by a tip in *P*. We define three types of missing cases: missing-in-tree (MIT), missing-off-tree (MOT), and completely missing (CM) based on the relationship with the observed phylogeny. The MIT cases are “in the tree” in the sense that some portion of the time that the pathogen was within that host corresponds to a portion of a branch in *P*. MOT cases are not represented on *P* directly, but their infection event can be traced back to an infectious period that is in *P*. And finally, the CM cases are those who are at least one generation removed from *P*. We distinguish two types of CM cases: the CM descendants and the externally CM. The CM descendants are cases who are descended from *P*, but whose infection event cannot be directly connected to it. Externally CM individuals are those whose most recent common ancestor with *P* lies outside *P* (in other words, they are not descendants of the root node of *P*, though they may be closely related). A concrete definition of these three types of missing is given below.

- **Missing-in-tree**. A case *i* is MIT if at least one of its descendants is sampled and *i* is descended from the root. For all MIT cases, some portion of their infection, from 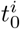 to 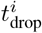, corresponds to some portion of *P*; the remainder of 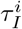 is dropped from the observed phylogeny. This time 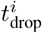 is defined as 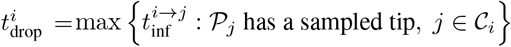.
- **Missing-off-tree**. A case *i* is MOT if none of its descendants are sampled and either
  - its infector is observed, or
  - its infector *j* is MIT and 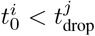.
- **Completely missing**. A case *i* is CM if none of its descendants are sampled and either
  - its infector is MOT or CM, or
  - its infector *j* is MIT and 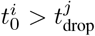.

The total number of MIT and MOT cases are denoted as *n*_MIT_ and *n*_MOT_, respectively. A diagram illustrating these three types of missingness is given in Fig 1. In this example, the phylogenetic tree for the whole outbreak is shown with solid edges indicating the portion of the phylogeny that is observable from the sampled sequences. In the example transmission graph, shown on the lefthand side of the figure, the MIT and MOT cases are directly connected to the observed infections while the CM case is not. Because of their relationship with the observed phylogeny, we can estimate the number of MIT and MOT cases; we refer to these cases as “cryptic”. We note that the number of cryptic cases is less than the number of unsampled cases, and should not be interpreted to be equal to the total number of unsampled cases. These will usually include the two kinds of cases we have called “completely missing”.

**Figure 1:**
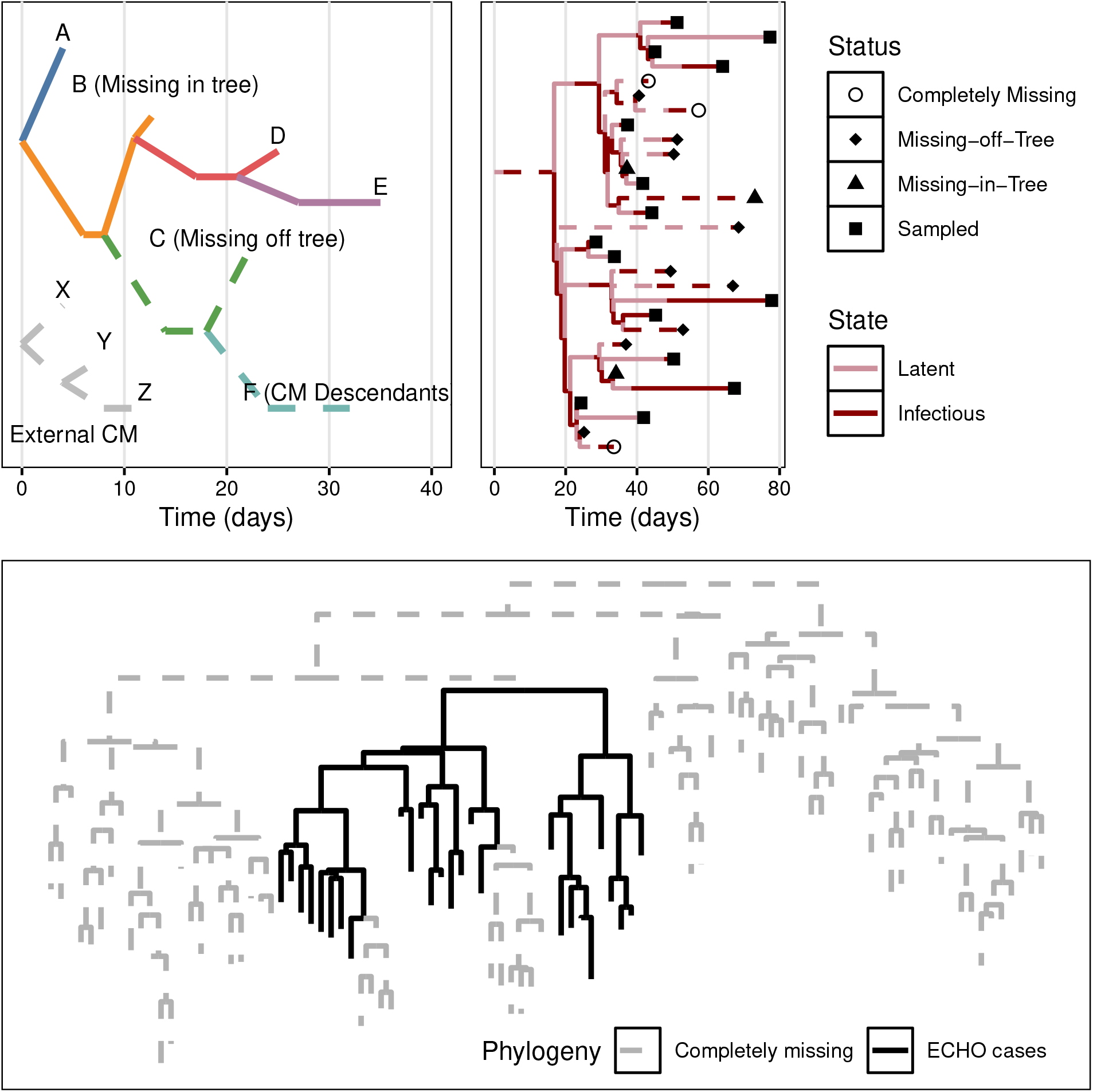
Missing case diagram. (Top left) A diagram of a partially observed transmission tree where the branch colour corresponds to the host. A, D, and E are sampled cases and the rest reflect the different types of missing cases. B is MIT because B infected D and D is sampled (so B is ancestral to the sample). We have information about B because of the time elapsed between A and D (both sampled). We have information about C because B was infectious for long enough that we expect C’s infection to have occurred. We do not extrapolate to estimate the numbers of CM descendants like F. (Top right) A diagram of a partially observed phylogenetic tree with edge colouring indicating the state of the infection. The tree comprised of the solid edges is *P*. Tip shapes indicate what type of missing each case is. The case counts in this example are *n*_obs_ = 10, *n*_MIT_ = 7, and *n*_MOT_ = 1. (Bottom) An example of how ECHO’s case count estimates relate to a broader outbreak. The tree in black contains all of the cases that are sequenced, MIT, or MOT and the cases in the tree in grey are completely missing.

### Estimators

In order to define a probabilistic relationship between cryptic cases and the observed phylogeny, we need a probabilistic model for infection progression. To this end, we limit our model to pathogens whose infection can be well modeled by a latent and infectious period. For an arbitrary individual *i*, we say that 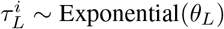, 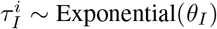, and 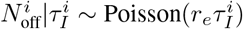, where *θ*_*L*_ and *θ*_*I*_ are the average duration of the latent and infectious periods, respectively, and *r*_*e*_ is the expected number of secondary cases generated per day. To be consistent with standard notation, we will specify the secondary case distribution through the effective reproduction number *R*_*e*_, the average number of secondary cases for an average individual, such that *r*_*e*_ = *R*_*e*_*/θ*_*I*_ . The three parameters needed to specify the disease progression model are represented by 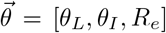. Since time on the phylogenetic tree is directly related to infection events between individuals, we can relate the total branch length on the tree to the total elapsed latent and infectious periods. Hence, the total tree length can be decomposed into portions corresponding to latent and infectious periods: | *P* | = | *P* |_*L*_ + | *P* |_*I*_ . From this we propose three relationships between total branch length and individual durations.

The first estimator (Estimator A) is based on solely the latent portion of the tree and is related to *n*_MIT_. To obtain an expression for | *P* |_*L*_, note that all observed and MIT individuals will have their full latent periods represented on the tree, so |*P*|_*L*_ will be the sum of these periods: 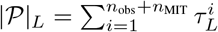. However, since only |*P*| is observed, we write | *P* |_*L*_ as | *P* | *×* (1 − *p*_*I*_ ), where *p*_*I*_ is the proportion of *P* that corresponds to the infectious stage. This unknown fraction can be approximated by

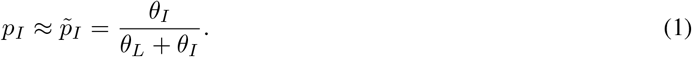

The expression in Eq 1 contains two nested approximations. The first approximates the proportion of the tree corresponding to the infectious period with the average proportion of their infection that an individual spends in the infectious period 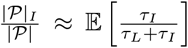. The second approximation brings the expectation operator inside the fraction, equating the expectation of the fraction with the fraction of the expectations, yielding the expression above: 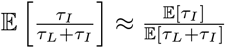. Numerical examination of these approximations has found them to be adequate for the settings tested here. Because | *P* |_*L*_ is a sum of independent and identically distributed exponential random variables, it has a gamma distribution:

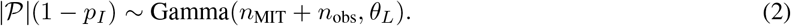

The second estimator (Estimator B) is also related to only *n*_MIT_, but uses the total branch length of the tree. This total branch length | *P* | includes the latent periods of all observed and MIT individuals, | *P* |_*L*_, plus the infectious periods for all observed individuals and the portion of the infectious period of MIT individuals up until *t*_drop_. This portion attributable to a MIT individual is *τ*_drop_ = *t*_drop_ − (*t*_0_ + *τ*_*L*_). The resulting expression for the total branch length is 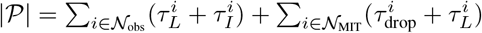. However, computing E[*τ*_drop_] would require specifying the sampling process. Instead, we observe that individuals with longer infectious periods and/or more secondary infections should be more likely to be MIT. Therefore, given that an individual is MIT, the average duration of their infectious period and their average number of secondary cases should be higher than of an arbitrary case. A derivation of these conditional quantities can be found in S2. Accordingly, we find that on average this conditional extension of the infectious period in and its truncation by *τ*_drop_ cancel out, leaving as a reasonable approximation 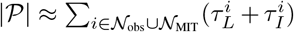. This results in |*P*| having a distribution defined by the sum of two gamma distributions with different scale parameters. While this does not have a simple closed-form distribution, we can approximate it with a gamma distribution whose scale parameter is the average of *θ*_*L*_ and *θ*_*I*_ :

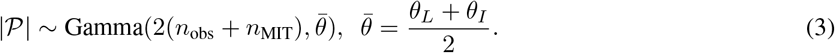

When *θ*_*L*_ and *θ*_*I*_ are similar to 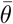, this approximation is reasonable and numerical examination is in agreement. Maximizing the resulting likelihood to obtain the shape parameter yields an estimate of 2(*n*_obs_ + *n*_MIT_). Given that *n*_obs_ is known, a simple transformation produces an estimate of *n*_obs_ + *n*_MIT_.

Finally, the third estimator (Estimator C) uses only the infectious portion of the tree and provides an estimate of *n*_obs_ + *n*_MIT_ + *n*_MOT_. To relate |*P*|_*I*_ to the number of cryptic cases we break the infectious periods into three segments: time to initial infection, the inter-infection intervals, and time to sampling. Say individual *i* infects individual *j* at time 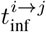, then individual *k* at time 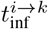, then is sampled at time 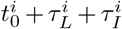. The time to first infection is the time from the onset of infectiousness until *i* infects *j*, 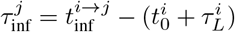. We also describe the elapsed time between subsequent infections, for example if *i* infects *j* and then *k*, we define their inter-infection intervals as 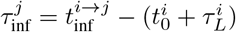 and 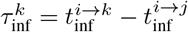. To complete the accounting of the infectious interval, we also need to introduce a 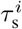, the portion of *i*’s infectious interval that lies between the final secondary infection (or the onset of their infectious period if they generate no secondary cases) and their sampling time. The transmission model implies that an individual will have *N*_off_ secondary cases that are distributed uniformly throughout an interval of length *τ*_*I*_ . Therefore, if the average individual generates *R*_*e*_ infections, their infectious period will be divided into *R*_*e*_ + 1 segments, so the expected values of the various *τ*_inf_ intervals, as well as *τ*_s_, are equal to 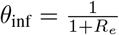. Therefore, the infectious portion of the tree can be written as 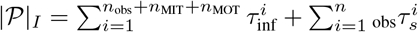. This gives us the distribution for the third estimator:

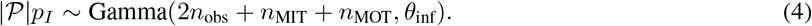

These three distributions are interpreted as likelihoods. The maximum likelihood estimates (MLEs) from the first two distributions give us an estimate of *n*_obs+MIT_ = *n*_obs_ + *n*_MIT_. The third distribution leads to an MLE of *n*_obs+MIT+MOT_ = *n*_obs_ + *n*_MIT_ + *n*_MOT_. The resulting estimators, A, B, and C, are summarized in Table 1.

**Table 1:**
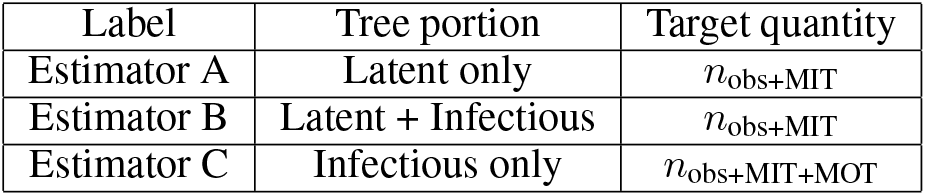
Summary of the three ECHO estimators.

| Label | Tree portion | Target quantity |
| --- | --- | --- |
| Estimator A | Latent only | $n_{\text{obs}+\text{MIT}}$ |
| Estimator B | Latent + Infectious | $n_{\text{obs}+\text{MIT}}$ |
| Estimator C | Infectious only | $n_{\text{obs}+\text{MIT}+\text{MOT}}$ |

To obtain confidence intervals for our estimates, we use a normal approximation. The typical asymptotic result from statistical theory is that the asymptotic variance of an MLE is equal to the inverse of its second derivative evaluated at the MLE. In the instance of a gamma distribution with shape parameter *n* the variance is [*ψ*^(1)^(*n*)]^−1^, where *ψ* is the trigamma function. The resulting (1 − *α*) *×* 100% CI for an estimate 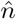 is

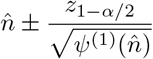

However, we advise caution when interpreting these intervals as the number of samples used to construct the observed phylogeny may be insufficient to support results from asymptotic MLE theory. The other source of uncertainty that can be taken into consideration is that about |*P*| arising from its construction from observed data. In a Bayesian analysis, often a collection of trees drawn from their posterior distribution is used to quantify the *a posteriori* uncertainty of *P*. This uncertainty can be propagated through ECHO by computing the ECHO estimates for each of the trees in the posterior sample and empirically estimating a 95% credible interval from them. This will capture uncertainty in the estimate arising from uncertainty in the total length of the tree, but will not capture the uncertainty from the individual MLEs.

All three ECHO estimation functions have been made available as an R package which can be installed from GitHub at https://github.com/MAGPIE-SFU/echo.

### Accuracy and empirical validation

We validate the performance of ECHO in several settings in a series of simulation experiments designed to evaluate the accuracy of ECHO, its sensitivity to the sampling scheme, and its sensitivity to model misspecification. Subsequently, we apply ECHO to trees reconstructed from measles virus (MeV) sequences from a published outbreak [12].

### Baseline simulation study

Artificial phylogenetic trees are simulated by first generating a transmission network from the model specified in the previous section. The simulation is run until *n*_max_ cases are generated, after which point the next generation of secondary cases is counted, but does not contribute to the case population. The phylogenetic tree corresponding to that network is reconstructed such that infection events are branching points and the sampled cases are the tips. From this complete phylogeny a sampled subtree is extracted by randomly sampling cases from the population. Each simulated dataset contains the total length of the observed phylogeny and *n*_obs_ as well as the true, unobserved values of *n*_MIT_ and *n*_MOT_.

The baseline simulation uses a parameter setting chosen to reflect measles in a partially vaccinated population: *θ*_*L*_ = 10, *θ*_*I*_ = 8, and *R*_*e*_ = 2 [13, 14, 15]. To establish a baseline for performance we use a fixed sampling fraction *ρ* at values *ρ* = 0.1, 0.2, …, 0.9. To evaluate the sensitivity of ECHO’s results to the sampling scheme across all three methods we compute the relative error, 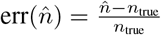.

### Robustness to heterogeneous sampling schemes

The term “phyletic group” is used here to refer to both monophyletic groups (or clades) and paraphyletic groups (a tree less some number of clades). We define *K* groups in a tree by randomly choosing *K* − 1 interior nodes; their descendants define *K* − 1 monophyletic groups. These clade roots are chosen such that the *K* − 1 clades are non-overlapping. We refer to the remaining part of the tree as “the paraphyletic group” (see Figure 3). To encourage reasonably-sized groups, the root nodes of the *K* − 1 monophyletic groups are sampled from cases whose infection time lies between 0.2*t*_max_ and 0.5*t*_max_. The goal of this experiment is to understand how differential sampling across groups affects the performance of ECHO, so we use *K* = 3 groups. To emulate different sampling schemes we fix a baseline sampling fraction *ρ* = 0.7 for the paraphyletic group; the sampling fractions for all three groups are defined by the composition ratios: 1:1:1 (all groups have sampling fraction 0.7), 1:1/2:1/2 (the paraphyletic group has sampling fraction 0.7 while the two monophyletic groups have sampling fraction 0.35), and 1:2/3:1/3 (the paraphyletic group has sampling fraction 0.7 and the remaining two groups have sampling fraction 0.47 and 0.23).

**Figure 2:**
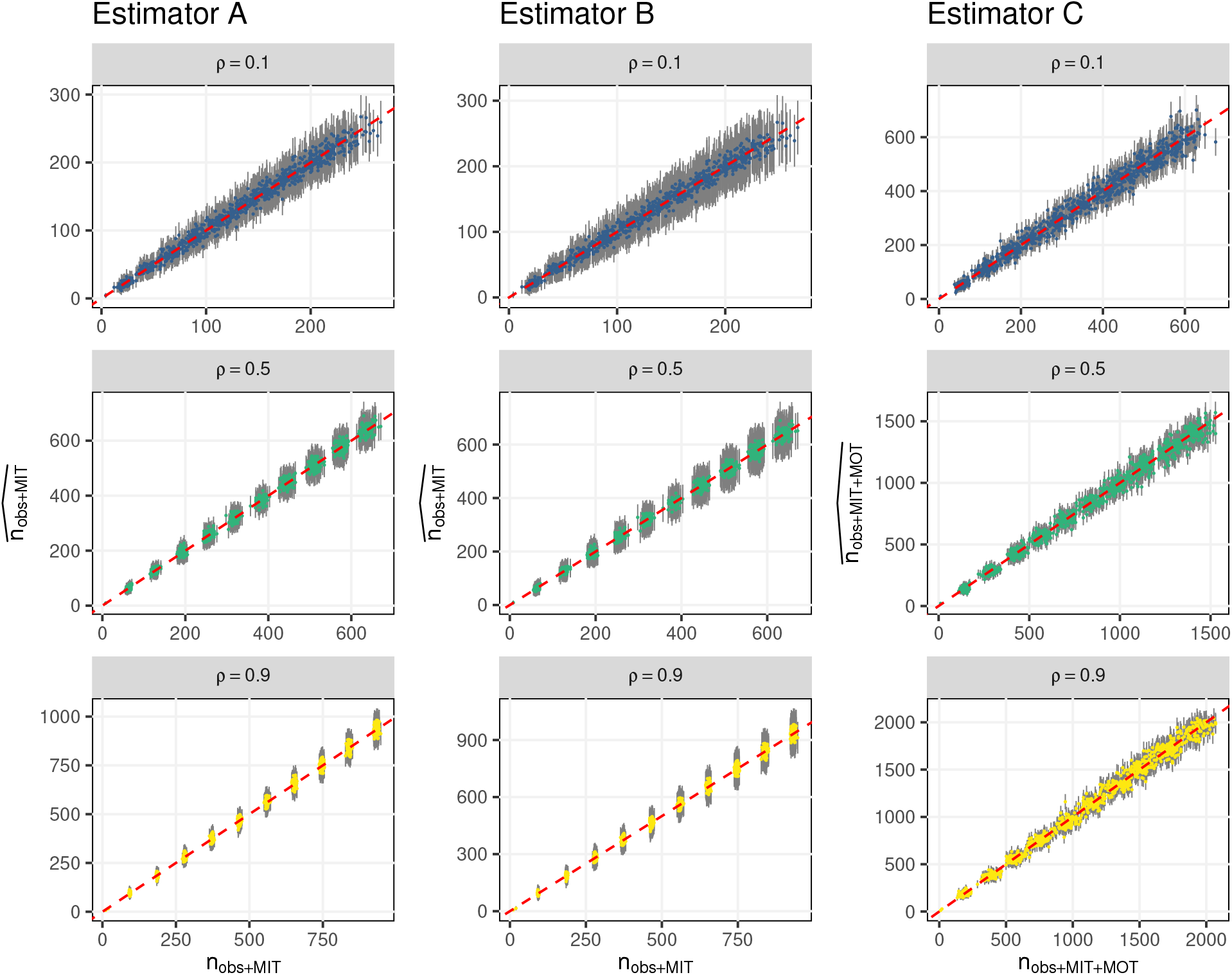
ECHO point estimates under fixed sampling fraction. Point estimates of *n*_obs+MIT_ (left and middle columns) and *n*_obs+MIT+MOT_ (right column) produced by the ECHO estimators. Vertical bars indicate the 95% confidence intervals. Input trees were generated with fixed sampling fractions of *ρ* = 0.1 (blue), 0.5 (turquoise), and 0.9 (green). The red dashed line is the axis of symmetry.

**Figure 3:**
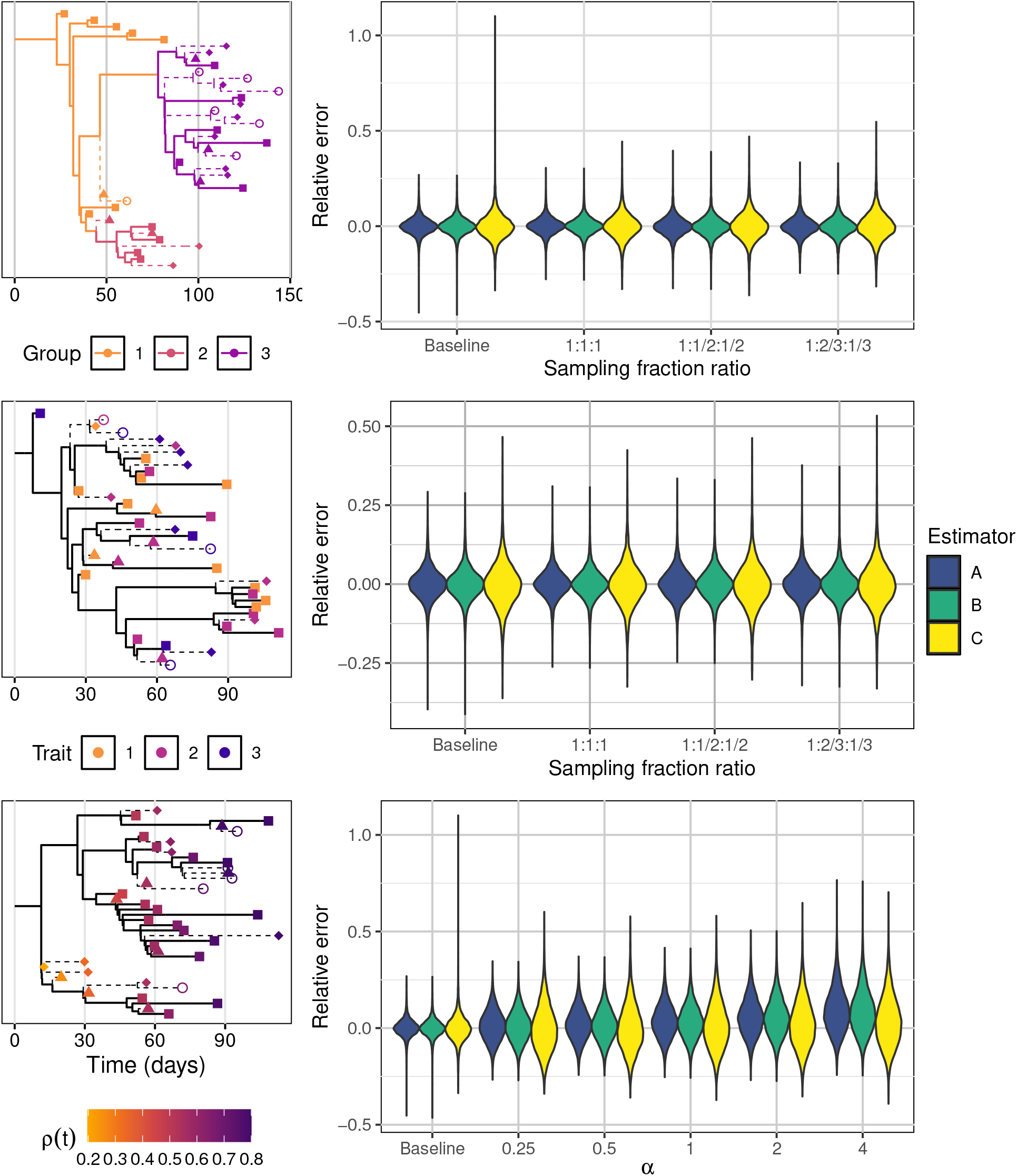
Error under heterogeneous sampling. (Top) A tree comprised of three groups with sampling fractions 0.70, 0.47, and 0.23 (a sampling fraction ratio of 1:2/3:1/3). (Middle) A tree with three traits whose sampling fractions are 0.70, 0.47, and 0.23. (Bottom) A tree with sampling rate increasing linearly through time (*α* = 1). The boxplots on the right show the relative errors for all of the trees in the corresponding experiment.

To attribute a discrete-valued trait to each case we use a continuous-time discrete-state Markov process with three states. Transitions between states are governed by an instantaneous transition rate matrix

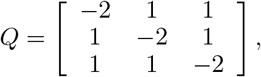

which implies that the transition between any two states are equally likely. The difficulty that arises in practice from trait-differentiated sampling is when one trait is disproportionately represented in the sample. Therefore we set a baseline sampling fraction, *ρ* = 0.7, and set three different trait compositions for the final sample. The composition of the sampling fractions between the three traits is defined in the same way as above for the three phyletic groups.

Time-varying sampling was examined through a function that increases with time. To do this we specify *ρ*(*t*), the probability of sampling a case at time *t* through a power law

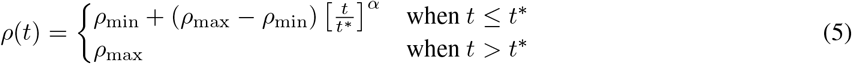

where *ρ*_min_ and *ρ*_max_ are the minimum and maximum sampling probabilities, *t*^*^ is the time at which the maximum sampling probability is reached, and *α* is a parameter that control the degree at which the sampling rate accelerates through time. In our experiments we set *ρ*_min_ = 0.1 and *ρ*_max_ = 0.8, so that *ρ*(*t*) covers a wide range. To account for the final generation of sampleable cases, we set *t*^*^ = *t*_max_ + *θ*_*L*_ + *θ*_*I*_ . The rate at which the sampling probability changes will be concave if *α <* 1, linear if *α* = 1, and convex if *α >* 1; we run our experiment at *α* = 1*/*4, 1*/*2, 1, 2, 4.

### Sensitivity to model and input misspecification

The next three experiments address the sensitivity of ECHO to model and input misspecification. The first two address parameter and distribution misspecification. The best distribution for modelling disease mechanics can vary widely based on the context of a particular outbreak, often driven by heterogeneity in the underlying population; establishing ECHO’s sensitivity to this type of misspecification is important for understanding its general utility. The transmission parameters are treated as fixed input, presumably chosen to reflect prior knowledge of the disease. In the third experiment we misspecify the total tree length |*P*| that is treated as observed by ECHO.

To evaluate the effect of rate misspecification we run ECHO with *θ*_*L*_ = 10, *θ*_*I*_ = 8, and *R*_*e*_ = 2 while varying the parameter values that generate the data. We assess the effect attributable to each parameter individually, varing each parameter separately while keeping the other two fixed at the assumed value; this design will ignore potential second and third order interaction effects between the parameters. Each parameter is varied at three levels: half the assumed value (*θ*_*L*_ = 5, *θ*_*I*_ = 4, *R*_*e*_ = 1), the assumed value (*θ*_*L*_ = 10, *θ*_*I*_ = 8, *R*_*e*_ = 2), and twice the assumed value (*θ*_*L*_ = 20, *θ*_*I*_ = 16, *R*_*e*_ = 4).

The only distributional assumptions behind ECHO are those of the disease progression model. All three distributions are one-parameter distributions whose mean determines its variance, a property which often does not arise in practice. Therefore, to model discrepancy between ECHO’s model and the actual data generative process we generate data from a gamma distribution for the latent and infectious intervals and a negative binomial distribution for the secondary case counts: distributions for which we can keep the mean fixed and vary the variance. We run three settings for each of the latent, infectious, and secondary case count distributions. The latent period distribution is parameterized by the mean *θ*_*L*_ and the variance scale *s* such that the variance is 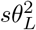; this results in a gamma distribution with shape 1*/s* and scale *sθ*_*L*_. A similar parameterization is used for the infectious period distribution. The secondary case count distribution is parameterized by the daily mean *r*_*e*_ and the dispersion parameter *k* such that the variance of *N*_off_ | *τ*_*I*_ is *τ*_*I*_ *r*_*e*_ + (*τ*_*I*_ *r*_*e*_)^2^*/k*. All three distributions are run under 5 settings, each run while leaving the other two distributions correctly specified. We use *s* = 0.25, 0.5, 1.25, 1.5, 2 for the latent and infectious periods and *k* = 0.1, 0.5, 1, 1.5, 2 for the secondary case counts.

Error in the value of |*P*| is emulated by first simulating *P* then adding or subtracting an amount proportional to |*P*| to the total branch length. For a tree with total length |*P*|, the value that ECHO uses to compute its estimates is (1 + *ϵ*)|*P*|, where *ϵ >* 0 is the proportional error in |*P*|. We use values of *ϵ* = −0.5, −0.25, −0.1, 0.25, 0.5, 1.0. As ECHO only uses the total branch length, error in the topology of *P* will not directly affect ECHO.

### Alternative outbreak settings

The preceding set of experiments all used values of 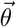 that were chosen to reflect measles in a partially vaccinated population. In the final two experiments we simulate outbreaks from different contexts; in both experiments ECHO is run with the correct parameter values. The first experiment emulates a measles infection in different vaccination/policy contexts. The parameters for the latent and infectious periods, *θ*_*L*_ and *θ*_*I*_, remain at the measles values used previously and *R*_*e*_ is varied to mimic the effect of differing context; values of *R*_*e*_ = 1, 2, 4, 6 are used. For each value of *R*_*e*_ we simulate trees with sampling fraction *ρ* = 0.1, 0.5, and 0.9.

In the second experiment we choose values for 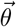 that are chosen to reflect other pathogens for which ECHO may be applicable. The first disease we consider is Ebola virus disease (Ebola). The evolution and transmission timeline of Ebola is suitable for the bottleneck model that ECHO relies on. Parameter values were taken from a review of epidemiological studies of Ebola: *θ*_*L*_ = 10, *θ*_*I*_ = 7, and *R*_*e*_ = 3 [16]. Since Ebola is able to transmit after the death of the host, we note that these parameter settings are chosen to reflect a setting in which sanitary funerary practices are adhered to reasonably well. The second disease is Mpox. In practice, the outbreak timeline for which Mpox is suitable for ECHO is longer than that of measles and Ebola due to the relatively slower rate of evolution, which affects the construction of the timed phylogenetic trees that ECHO uses.

All experiments are run on 10000 simulated trees under each experiment setting. These trees are run with *n*_max_ = 100, 200, …, 1000 (1000 trees in each setting). Experiments are run with a sampling fraction of *ρ* = 0.5, unless stated otherwise. To evaluate the effect of the experiment setting on the error of the ECHO estimators, the relative errors are plotted alongside a baseline error. As a baseline we use the results from the experiment with a uniform sampling fraction. All simulations are run in Julia [17] and can be replicated at https://github.com/MAGPIE-SFU/echo-experiments; all plots are generated in R [18].

### Application to measles virus

We apply ECHO to MeV sequences collected during an outbreak in the USA in late 2021 [12]. During the resettlement of Afghan evacuees in 2021, measles was detected among the evacuating population. In the three months following the initial detected cases, evacuees entering the USA were monitored for symptoms, quarantined, and in many cases, viruses from infected individuals were sampled and sequenced. Due to the highly controlled nature of this outbreak, the total number of infected individuals is known with a high degree of certainty, making this dataset a good test case for ECHO. We replicate the original analysis in BEAST2 [19] using the XML file provided by the authors. The original analysis used a skyline coalescent tree prior, a TIM+F+G4 substitution model for the coding regions, and a TIM3+F+G4 for the non-coding regions. A more detailed description of the model and its justification can be found in the original paper. This analysis was left unchanged except for the mutation rate, which we replaced with a fixed value of 7.5 *×* 10^−4^, to more closely align with estimates from the literature [20]. In addition to the phylogenetic analysis, Masters et al. [12] estimated parameter values of *θ*_*L*_ = 8, *θ*_*I*_ = 5, and *R*_*e*_ ≈ 2, which we use as inputs for ECHO. Since we replicate the BEAST2 analysis, we compute ECHO estimates for a posterior sample of phylogenetic trees. This analysis is conducted in R.

## Results

### Simulation study

The point estimates produced by each of the ECHO estimators are shown in Fig 2 for *ρ* = 0.1, 0.5, and 0.9. All three methods are reasonably accurate on average. An examination of the relative errors (Fig S1 in S1) shows nearly all simulated estimates being within 50% of the true value and most within 25%. The 95% confidence intervals (indicated by the vertical bars) tend to capture the true value, however it appears to overestimate the uncertainty when the estimated quantity is lower. There is no clear evidence suggesting that the performance of ECHO is affected by the sampling fraction.

The accuracy of ECHO is mostly unaffected by heterogeneous sampling, as shown in Fig 3. An example of each type of sampling process is shown on the left. The group- and trait-based sampling processes have sampling fractions of 0.70, 0.47, and 0.23; the time-based sampling uses *α* = 1 (a linearly increasing sampling probability). The presence of sampling heterogeneity across phyletic groups or discrete traits has very little effect on the accuracy of ECHO. The results for time-based sampling are similar, except with some effect on Estimator B when the sampling increases sharply with time (large *α*). Recall that Estimator B uses an approximation that relies on mostly uniform sampling within individual infectious periods – this may not be the case when the sampling probability increases rapidly. Robustness to model misspecification varies. Misspecifying the average duration of the latent or infectious periods has a roughly proportional impact on accuracy of all three estimators while only Estimator C is sensitive to *R*_*e*_ (see Fig S5 in S1). The variance of the distributions (see Fig S6 in S1). Over- or underdispersion of the latent or infectious period does not affect the accuracy of any of the ECHO estimators. Similarly, overdispersion of the secondary case counts also has no effect on Estimators A and B. However, at relatively higher levels of overdispersion, Estimator C tends to underestimate *n*_obs+MIT+MOT_.

The greatest determinant of error in ECHO was discrepancy between the total length of the input tree and the true phylogeny (see Fig S7 in S1). We observe a roughly linear relationship between the error in |*P*| and the error of the ECHO estimators. In the most extreme case, when the observed |*P*| was twice that of the true tree, the all three estimators tend to overestimate their respective quantities by the same factor. ECHO also performs well on simulations with different parameters from the baseline (see Supplementary Materials).

### Measles application

The original analysis of the MeV sequences from the OAW outbreak identified three clusters of epidemiologically linked cases that were associated with distinct introductions. Those clusters contained 32, 8, and 1 individuals, respectively. Additionally, the authors identified 47 total cases associated with this outbreak with no evidence of broader community spread. We focus on the first two clusters. The maximum clade credibility tree for each cluster and the histogram of the results from ECHO for all trees in the posterior sample generated by BEAST2.5 are shown in Fig 4. Using Estimator A we estimated a median (across all posterior samples) of 38 cases (6 MIT) associated with the first cluster and 8.2 (0.2 MIT) cases with the second cluster. This corresponds to a total of 46.2 total cryptic cases, which is 0.2 away from what we were expecting to find.

**Figure 4:**
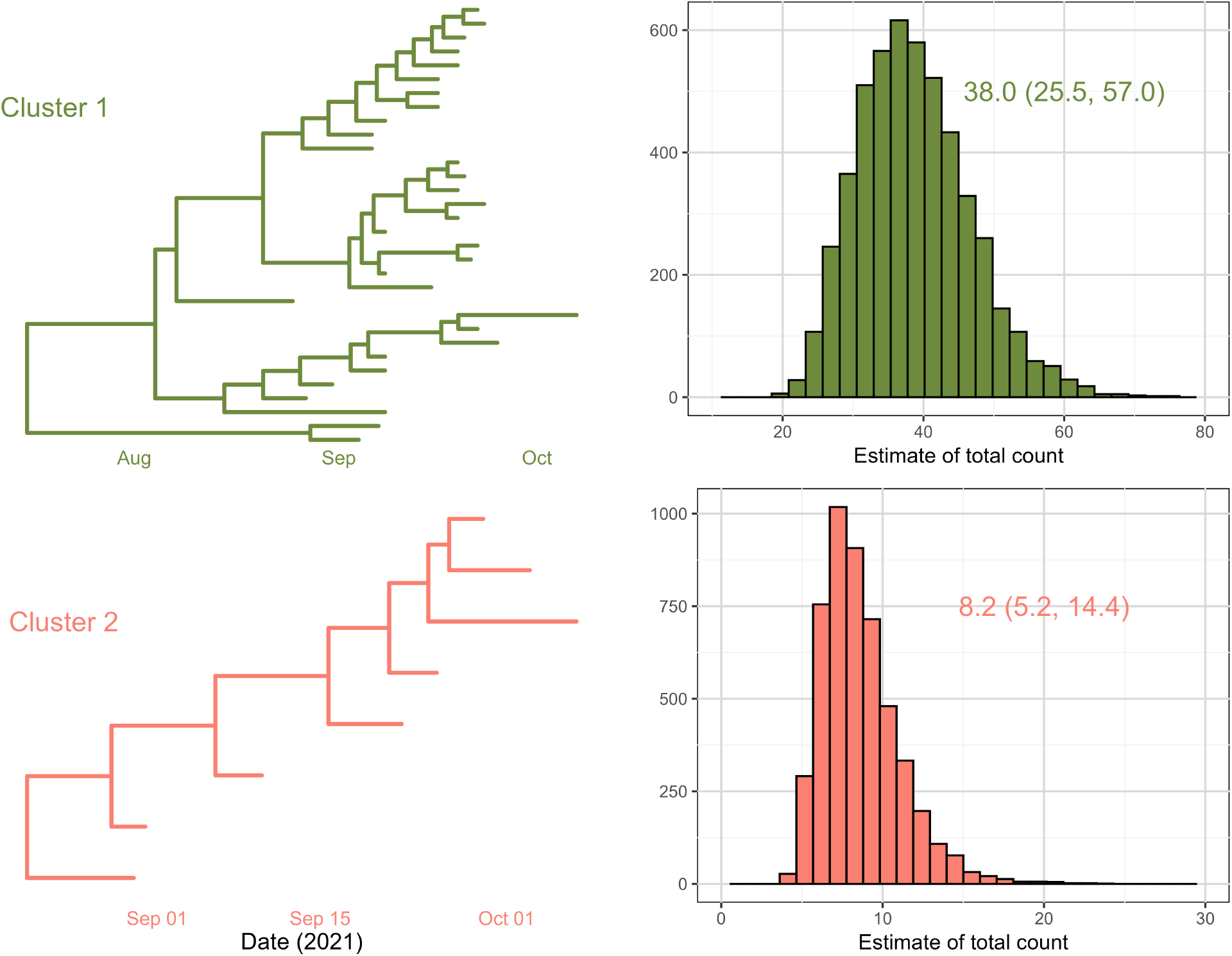
Inference results for MeV application. Maximum clade credibility trees for two MeV clusters and the associated results from ECHO. The histograms show the point estimates computed by ECHO for each of the trees in a posterior sample. Annotations indicate the point estimate and 95% credible interval produced by Estimator A.

Table 2 gives the point estimates and 95% credible intervals for all three of the ECHO estimators, for both the individual clusters and the combined data. Estimators A and B, which both estimate *n*_obs+MIT_, provide similar estimates, each well within the 95% credible intervals of one another. Estimator C produces much higher estimates because it is estimating *n*_obs+MIT+MOT_, which includes an additional generation of cases. We compare the estimates of *n*_obs+MIT_ to the true value taken from the study because the quarantine and vaccination strategy that was adopted to control the outbreak was successful and therefore the final generation of infections likely did not produce any secondary infections. Furthermore, because of the nature of the relevant population and the intensity of measles control efforts, it is likely that there were few completely missing cases.

**Table 2:** Estimates of total case counts for the two clusters in the MeV example.

| Method | Cluster 1 | Cluster 2 | Combined |
| --- | --- | --- | --- |
| Estimator A ($n_{\text{obs}+\text{MIT}}$) | 38.0 (26.0, 50.0) | 8.20 (2.8, 13.7) | 46.0 (32.8, 59.3) |
| Estimator B ($n_{\text{obs}+\text{MIT}}$) | 39.1 (23.7, 62.6) | 7.92 (4.24, 15.7) | 47.4 (30.4, 72.4) |
| Estimator C ($n_{\text{obs}+\text{MIT}+\text{MOT}}$) | 113 (75.6, 170) | 23.6 (14.8, 42.4) | 137 (96.4, 198) |

Point estimates are the posterior median and the brackets indicate the 95% credible interval of the point estimate.

### Interpretation

In many contexts it is unlikely to be the case that a very high fraction of infections are detected through testing (or in particular that a high fraction are sequenced, as in the measles example). In this case, ECHO’s estimates of the numbers of cryptic cases ars different from the total number of cases. It may be possible to extrapolate from ECHO’s estimates, although this requires additional assumptions and would not preserve ECHO’s robustness to variation in sampling across the tree or over time. For example, consider a branching process in which individuals are sampled with probability *π*, and in which the distribution of the number of secondary infections has probabilities *p*_*k*_ for *k* secondary infections and probability generating function *g*(*s*). In this case the long-time probability *ω* that an individual is unsampled, and all infections descending (at any generation) from the individual are also unsampled, satisfies the well-known equation

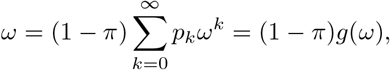

which can readily be solved numerically and which can be adapted to account for finite time [8]. The probability of being ancestral to the sample is 1 − *ω*. The sampling probability *π* is unknown, but ECHO gives the probability *p*_*E*_ of being sequenced given being ancestral to the sample (ECHO computes the number of missing-in-tree cases, and the number of sequenced cases is known). This results in two equations, *π* = *p*_*E*_(1 − *ω*) and the above, which can be solved simultaneously. This in principle can give rise to an estimate of the total number of infections, 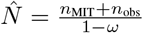. However, computing *ω* requires either the long-time assumption or averaging over unknown times of infection. Similarly, in a continuous time branching process with constant birth, death and sampling rates, the probability of a lineage becoming extinct (similar to *ω*), *E*(*t*), is the solution to a Riccati equation, *dE/dt* = *µ* − (*λ*+*µ*+*ψ*)*E* +*λE*^2^ [21] with boundary condition reflecting sampling assumptions. This can be extended to state-specific extinction probabilities [22] if birth and death rates vary across discrete states, and could in principle be used in the same way as *ω* with the same caveats.

## Discussion

We present ECHO, a simple and robust tool for estimating the the total number of cases related to a sample of pathogen genomes. As sequencing technology becomes more affordable, phylogenetic analysis is becoming a standard tool used to manage pathogen outbreaks. While there are many methods that estimate the parameters of epidemiological and evolutionary models, the sampling fraction often must either be known *a priori* or follow a known process, which can be limiting in many practical settings. In contrast, ECHO connects the total phylogenetic time of an outbreak, estimated by timed tree reconstruction methods, to the number of cryptic cases through known information about the epidemiology of a disease.

The extent to which a case is missing is complex in the context of phylogenetics. We distinguish between missing-in-tree cases, those who occupy some portion of the observed phylogeny; missing-off-tree cases, those whose infection event can be directly linked to the observed tree; and completely missing. We can further distinguish the completely missing cases into those who are completely missing descendants of the observed phylogeny, *e*.*g*., the offspring of a MOT individual, and those who are completely missing and external to the tree, *i*.*e*., not a descendant of the tree’s root. Because the MIT and MOT cases are directly related to the observed phylogeny, they represent a weaker state of missingness than the completely missing cases. By making a small number of reasonable assumptions, ECHO is able to accurately estimate the number of these ‘cryptic’ cases. However, ECHO’s estimates should not be interpreted as the total number of unsampled cases, because they do not go beyond cases either ancestral to the sample or infected directly by an infection that is ancestral to the sample. If the number of sequences is small compared to the size of the overall outbreak, ECHO’s estimates will not reflect the total outbreak size.

There are many methods that use genomic data with phylogenetic models to infer molecular clock rates or epidemiological parameters. ECHO, however, makes use of existing knowledge of these quantities to produce three simple estimators. One of the benefits of this simplicity is that we avoid re-estimation of well-established quantities. This means that ECHO can be incorporated into existing genomics workflows without requiring specialized software. The other quality arising from ECHO’s simplicity is its robustness to heterogeneity in the sampling process. This phenomenon of heterogeneous sampling frequently occurs in realistic data settings as sampling fractions can vary by health jurisdiction, demographics, time, or other determinants. While several existing methods are able to accommodate complex sampling processes, they are often sensitive to having that process specified accurately. However, as ECHO is agnostic to the sampling process, performance does not depend on its form. Additionally, in many outbreaks there are multiple introductions from epidemiologically distinct sources. When this happens, the most recent common ancestor of all cases lies outside of the outbreak and the branch lengths will represent cases that are not associated with the population of interest. Multiple introductions can be accommodated by ECHO by simply providing the total branch length across all of the clades arising from these distinct introductions. This functionality was demonstrated in our application to MeV data. In this MeV analysis, we also demonstrated how ECHO can be incorporated into Bayesian analysis pipelines. A sample of trees from a posterior distribution can be fed tree-by-tree into ECHO to obtain a posterior distribution over the number of cryptic cases. Since ECHO is computationally lightweight this will be a negligible addition to the overall computation time, when compared to the runtime of most Bayesian phylogenetic software.

The simplicity of ECHO does impose limitations as well. As ECHO requires only four inputs, its results are somewhat sensitive to those inputs. In particular, misspecification of the length of the input phylogeny and some of the epidemiological parameters (*θ*_*L*_ and *θ*_*I*_ ) is detrimental. However, only Estimator C is affected by *R*_*e*_. The risk of estimation error can be offset by incorporating input uncertainty into ECHO. While this is not a current feature of the software, modifying the ECHO estimators to account for *a priori* uncertainty about its inputs by creating a Bayesian version of ECHO would be straightforward. Another limitation of ECHO that is of particular relevance to practical application is that we assume that the end of infectiousness is coincidental with sampling. This will not be a realistic assumption in scenarios where transmission often continues for a substantial time after sequencing. ECHO also does not take within-host evolution into account, as it uses the common phylodynamic assumption that branching events in the phylogeny correspond to transmission events. Within-host evolution would add additional length to the phylogeny; accounting for that may be a useful extension if the necessary information about within-host evolution is available.

While ECHO is applicable in many settings, there are certain pathogens or outbreak settings for which it is not well suited. ECHO requires a reasonably accurate timed phylogenetic tree (at least in terms of total length) and knowledge of the latent and infectious periods. Constructing an accurate phylogenetic tree requires a sufficient amount of genetic variation between samples. In general, this means that rate of evolution of a pathogen determines the sampling density suitable for phylogenetic reconstruction [23]. This, in turn, imposes a requirement on the nature of the sampling, rather than on the pathogens themselves. For RNA viruses (e.g., measles, Ebola) and fast-evolving bacteria, ECHO can be applied to outbreaks with fairly dense sampling. On the other hand, for DNA viruses (e.g., Mpox) and slowly evolving bacteria, ECHO can only be applied to data sampled sparsely enough to ensure sufficient genetic variation to create timed phylogenetic trees.

The latent/infectious model used by ECHO imposes more directly a restriction on the pathogens that are compatible with ECHO. Pathogens and diseases for which this latent/infectious model is inappropriate will be inappropriate for ECHO. This includes vector-borne illnesses (e.g., malaria, dengue) and pathogens with primarily zoonotic transmission (e.g., H5N1 and other zoonotic influenza). While it may be possible to extend ECHO to accommodate multiple host species, at present these pathogens are incompatible with ECHO. A high degree of variability in the duration of the latent or infectious intervals also is not consistent with ECHO’s underlying model. Tuberculosis, for example, has a long and highly variable latent period, making it unsuitable for ECHO.

Data privacy is a concern for public health agencies in many regions. The importance of genomic data to the management of outbreaks is offset by the sensitive nature of its privacy concerns. Even a reconstructed phylogenetic tree may be impermissible to transfer between research groups without explicit ethics approval or censoring of the tip dates. While a phylogenetic tree may contain identifiable information, its total branch length contains no information about any underlying individuals. Because ECHO requires only the total branch length of the tree, the relevant information can be shared between groups without any of the identifying information.

In relating a model of infection that includes latency to phylogenetic trees, we identify a conflict between phylogenetic models and epidemiological models. While an infection is latent, the pathogen is unable to transmit and is undetectable by most tests. In the context of a pathogen phylogeny, this means that branching/coalescent events cannot happen during a latent period. However, the standard phylogenetic models do not impose any restriction on branch lengths and as a consequence, in practice the inferred phylogenetic trees can have branch lengths that are inconsistent with a model of latency. In this paper we do not propose a resolution to this conflict, however the results of ECHO should be interpreted cautiously when the phylogenetic model produces trees that are inconsistent with pathogens with known latency.

## Data Availability

Only simulated and previously published data were used in this study. The previously published measles sequences can be found online at: https://data.cdc.gov/Models/Measles-Case-and-Genetic-Metadata-Operation-Allies/b8tp-jsmh/about_data

## S1 Supplemental Figures

Here we provide a guide to the supplementary figures. Each figure shows the relative error of the three ECHO estimators; full context can be found in the main manuscript.

S1 **Fixed sampling fraction for settings reflecting measles, Ebola, and Mpox**. The sampling fraction is varied from 0.1 to 0.9 and three disease settings are considered: measles (*θ*_*L*_ = 10, *θ*_*I*_ = 8, *R*_*e*_ = 2), Ebola (*θ*_*L*_ = 8, *θ*_*I*_ = 7, *R*_*e*_ = 3), and Mpox (*θ*_*L*_ = 8.5, *θ*_*I*_ = 14, *R*_*e*_ = 2.5).

S2 **The sampling fraction varies by phyletic group**. The tree is partitioned into three phyletic groups, each of which has its own sampling fraction. With a baseline sampling fraction of 0.7, we consider three settings determined by the following ratios: 1:1:1, 1:1/2:1/2, and 1:2/3:1/3.

S3 **The sampling fraction varies by a discrete-valued trait**. Traits were assigned to each case based on a continuous-time Markov process. The transition matrix was chosen such that all three traits are equally likely at stationarity. The sampling fractions were set such that one trait had a sampling fraction of 0.7 and the other two were determined by the following ratios: 1:1:1, 1:1/2:1/2, and 1:2/3:1/3.

S4 **The sampling probability is a function of time**. The sampling probability of a case is based on the time at which they were sampled. The rate at which the sampling probability increases is determined by the parameter *α*. The form of the sampling probability function is given in the main manuscript and is illustrated in the figure for different values of *α*.

S5 **Misspecification of the three epidemiological parameters**. We run ECHO’s inferences with assumed values of *θ*_*L*_ = 10, *θ*_*I*_ = 8, and *R*_*e*_ = 2 and change the values from which we generate the data. The experiment is run under three settings of parameter specification: correctly specified (*θ*_*L*_ = 10, *θ*_*I*_ = 8, *R*_*e*_ = 2), data generated with values lower than are assumed in inference (*θ*_*L*_ = 5, *θ*_*I*_ = 4, *R*_*e*_ = 1), and data generated with values higher than assumed in inference (*θ*_*L*_ = 20, *θ*_*I*_ = 16, *R*_*e*_ = 4). For each specification setting, one parameter is varied while the other two are held at the correct value.

S6 **Misspecification of the distribution families in ECHO’s epidemiological model**. We use a gamma distribution for the duration of the latent and infectious periods and a negative binomial distribution for the secondary case count distribution. We use the same value for the mean between ECHO and the simulator and vary the parameter that controls the variance for the gamma/negative binomial distributions.

S7 **Incorrect total branch length on the input tree**. We emulate the effect of error in the total branch length of the input phylogeny by simulating trees and adding or subtracting an amount proportional to their total branch length. We consider error rates from 0.5 (50% too short) to 1.0 (100% too long).

S8 **Variation of both the sampling fraction and effective reproduction number**. In these experiments ECHO uses the correct value. This experiment validates that the accuracy of ECHO’s approximations, evidenced in previous Figures, were not an artefact of a particular choice of *R*_*e*_.

**Figure S1:**
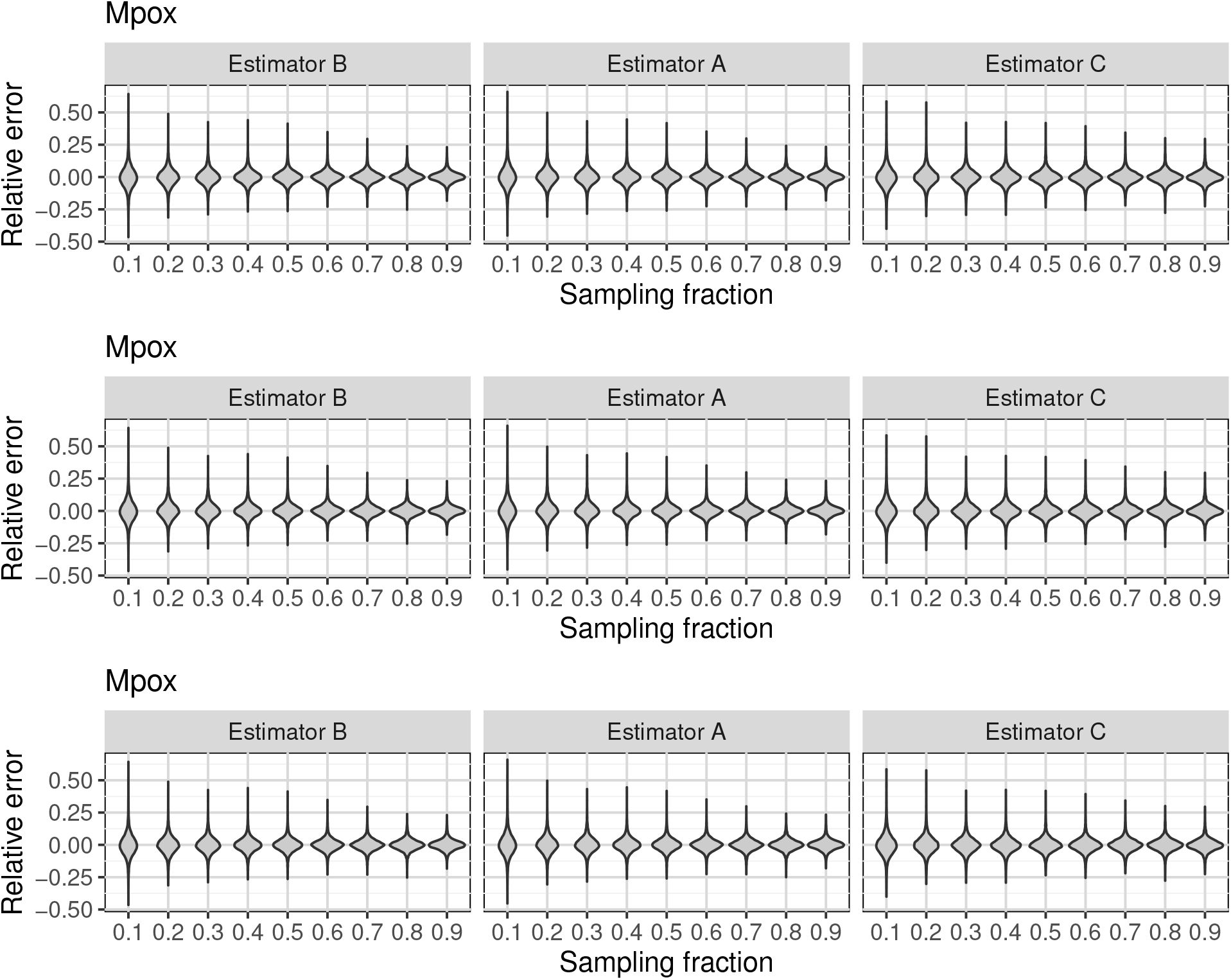
Relative error of the ECHO estimators under settings for measles (top) Ebola (middle) and mpox (bottom).

**Figure S2:**
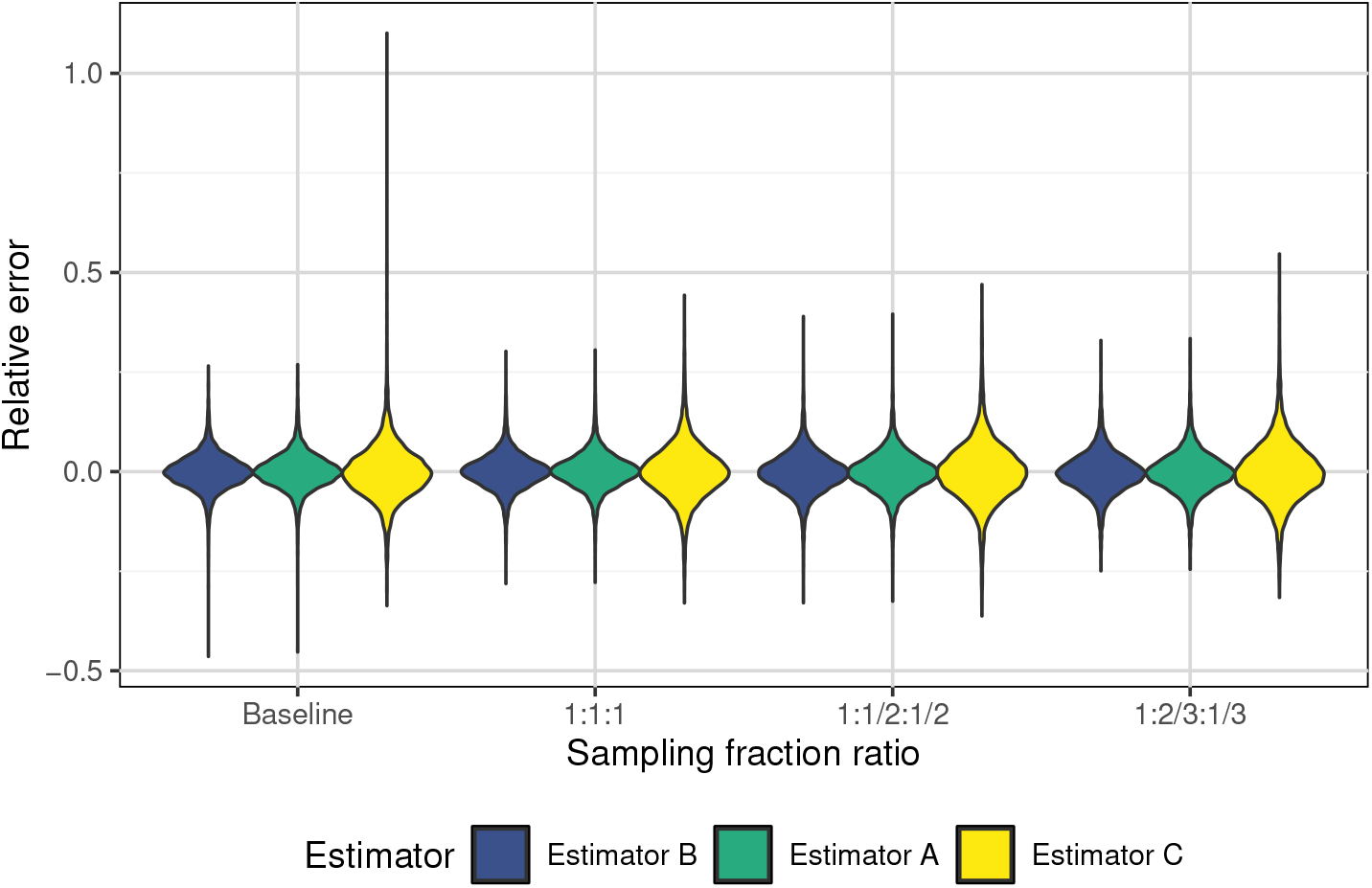
Accuracy of ECHO when the sampling fraction varies across phyletic groups. The sampling fraction ratio describes the sampling fractions of the groups, relative to a baseline fraction of 0.7.

**Figure S3:**
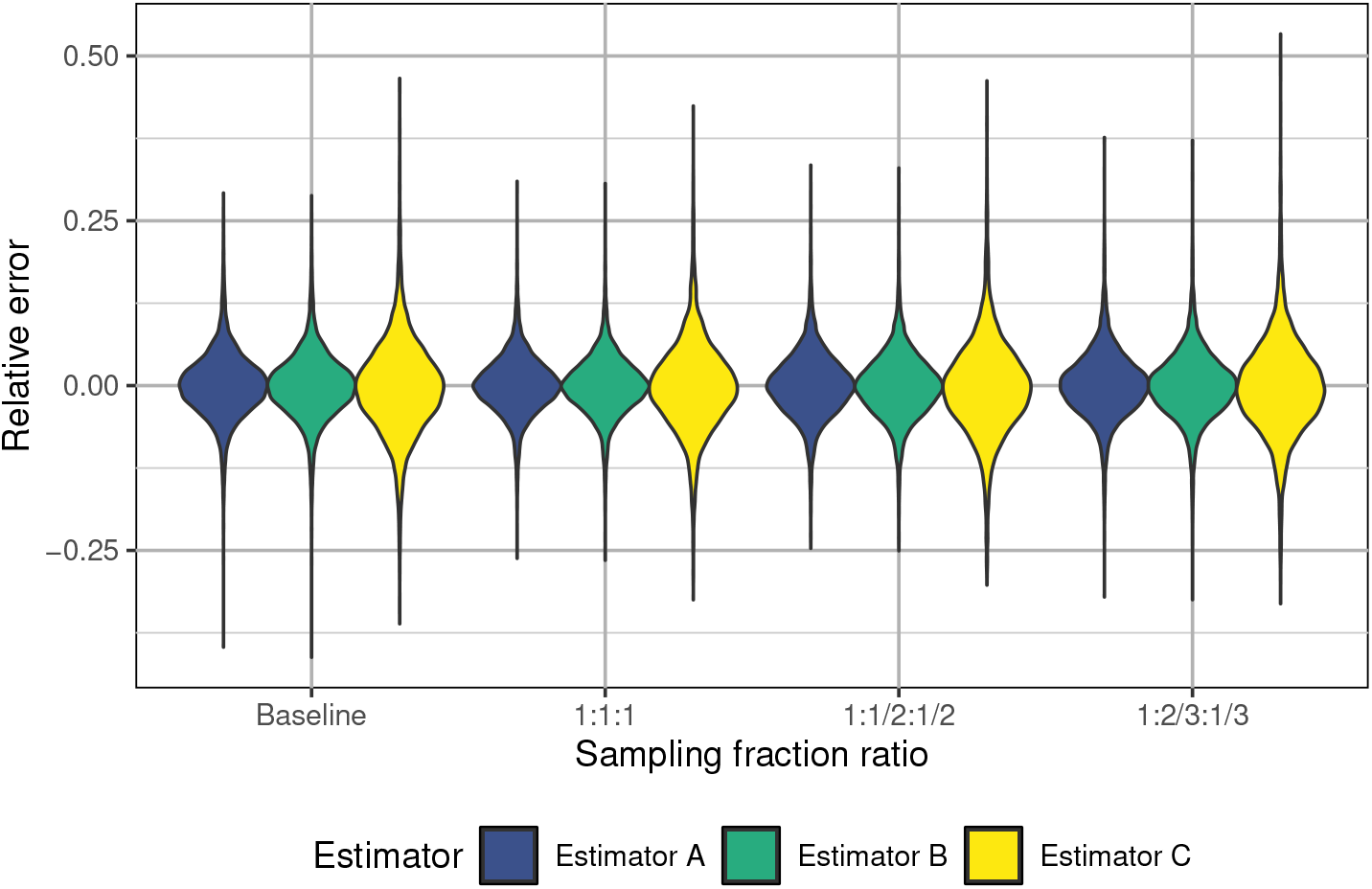
Relative error when the sampling fraction varies by a discrete-valued trait of the case. The sampling fraction ratio determines the relative sampling fractions across the three traits, relative to a sampling fraction of 0.7.

**Figure S4:**
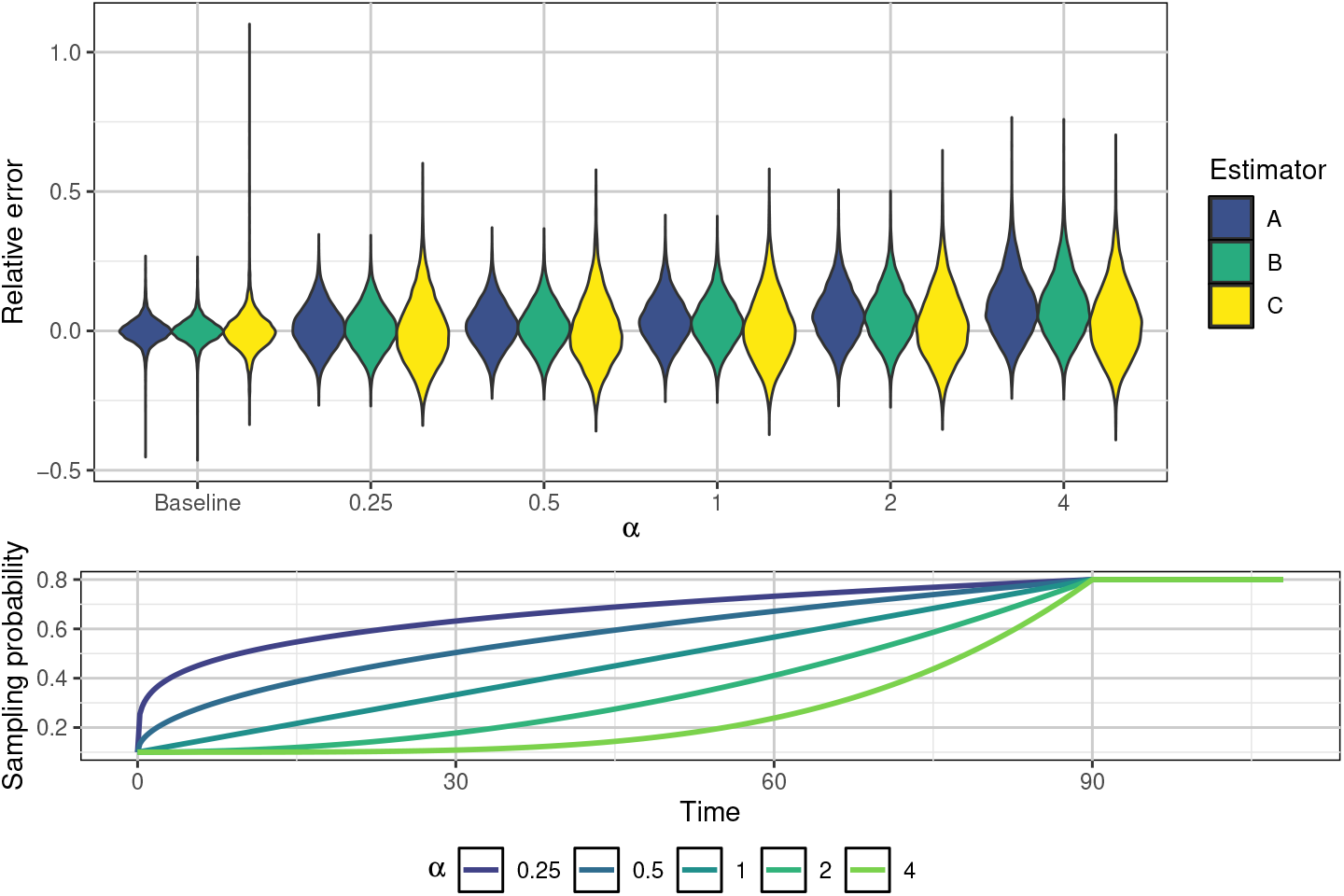
(Top) Relative error when the sampling probability is a function of time. The parameter *α* determines the shape of the sampling function as defined in the manuscript. (Bottom) The sampling probability as a function of time under each value of *α*.

**Figure S5:**
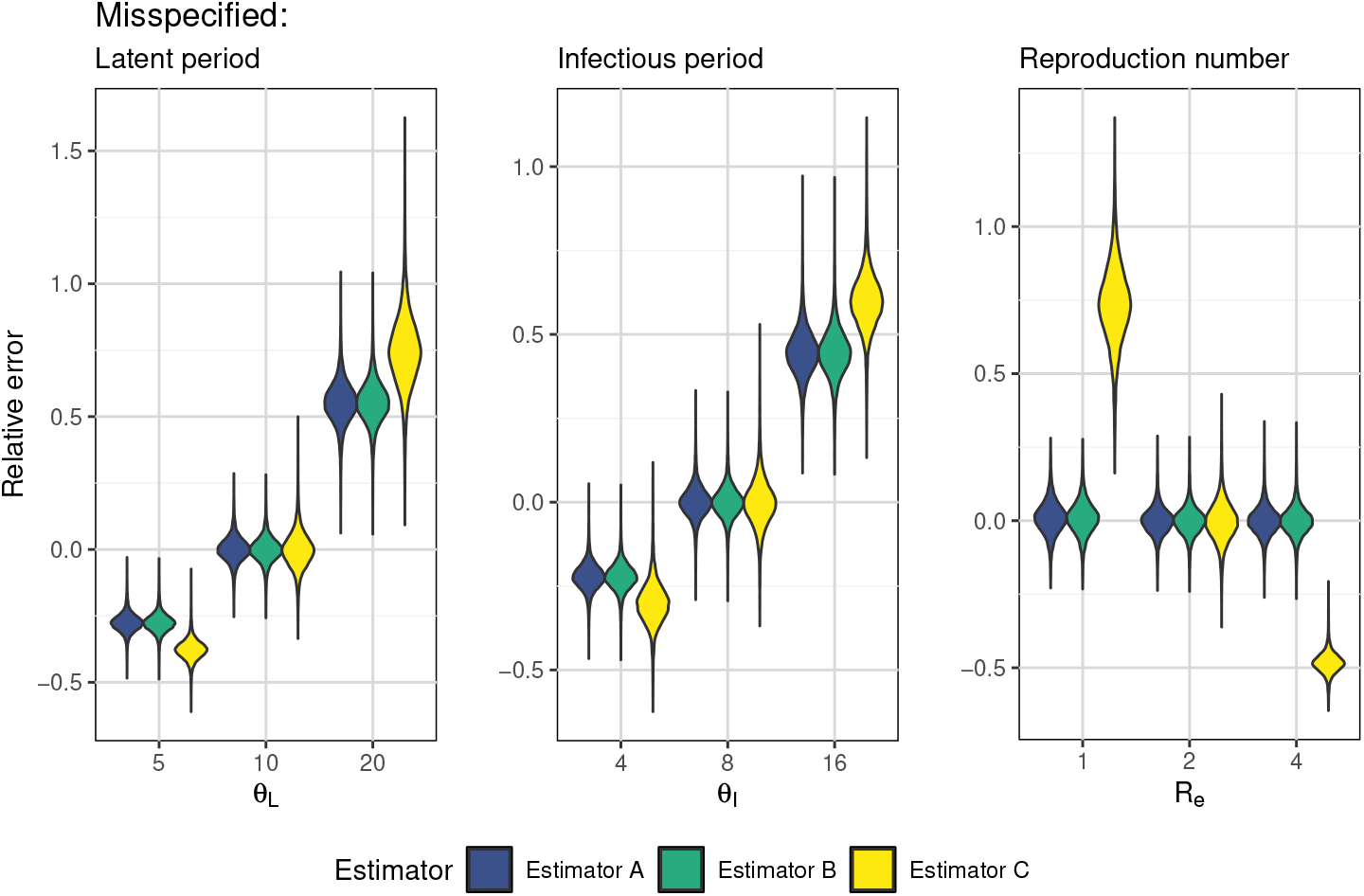
Relative error when the three epidemiological parameters that are used in ECHO are misspecified. The values on the *x*-axis are the values that are used to simulate the trees; ECHO is run using *θ*_*L*_ = 10, *θ*_*I*_ = 8, and *R*_*e*_ = 2.

**Figure S6:**
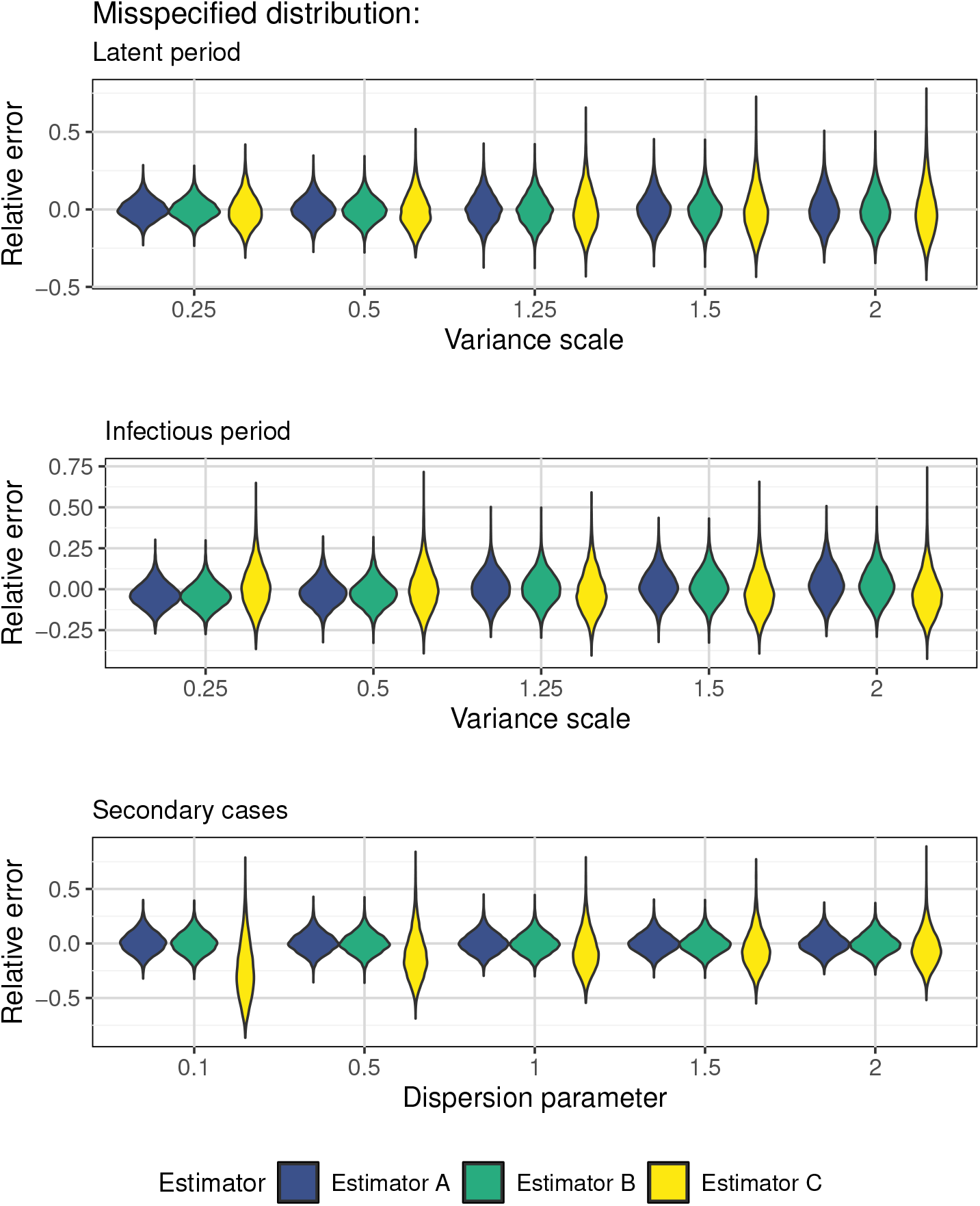
Relative error when the distributional family of the stochastic epidemiological model used by ECHO is incorrect. Using gamma distributions for the latent and infectious period and a negative binomial distribution for the secondary case counts, the parameter controlling the variance is on the *x*-axis.

**Figure S7:**
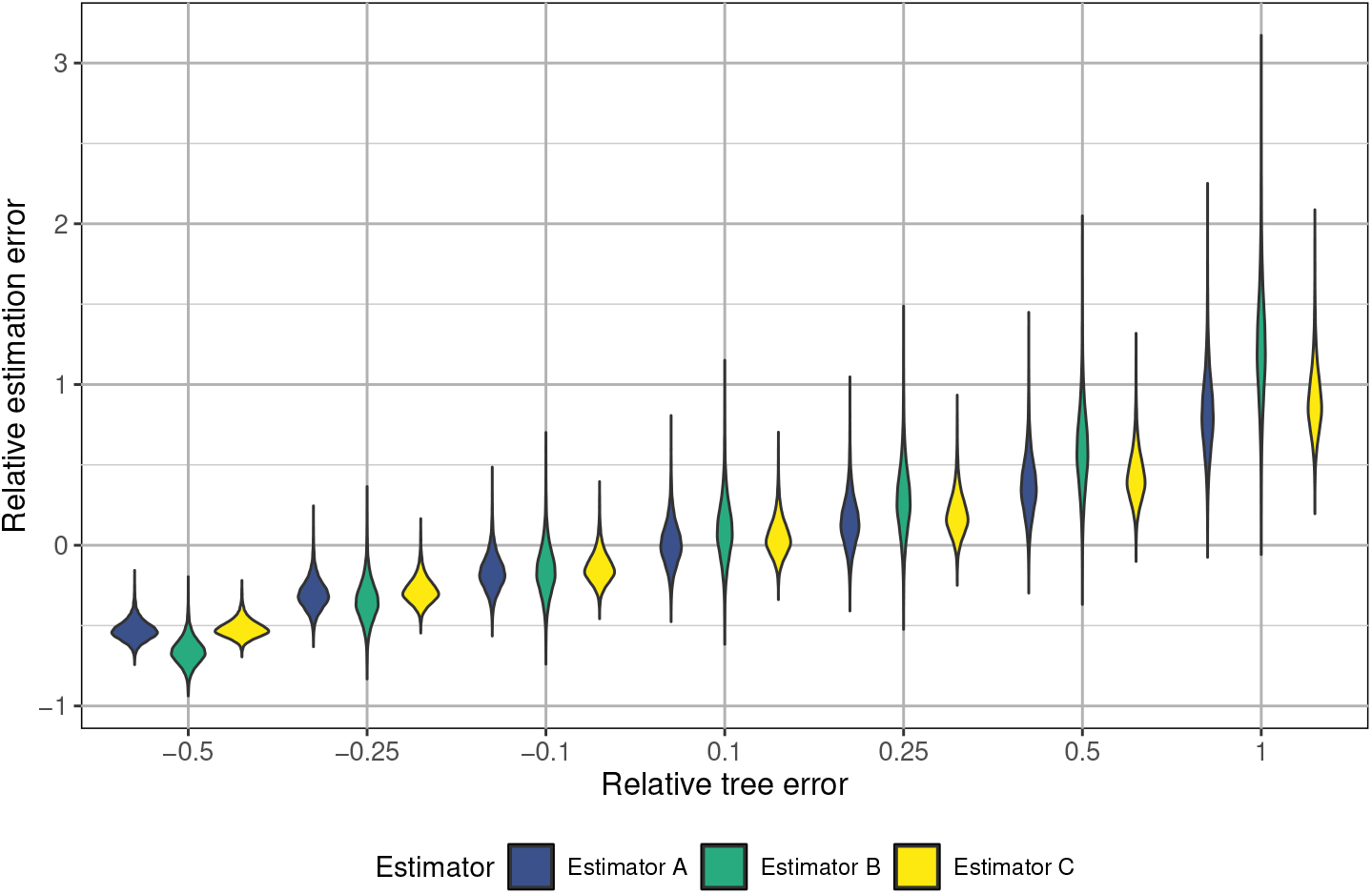
Relative error when the input tree is incorrect by an amount proportional to its true total branch length.

**Figure S8:**
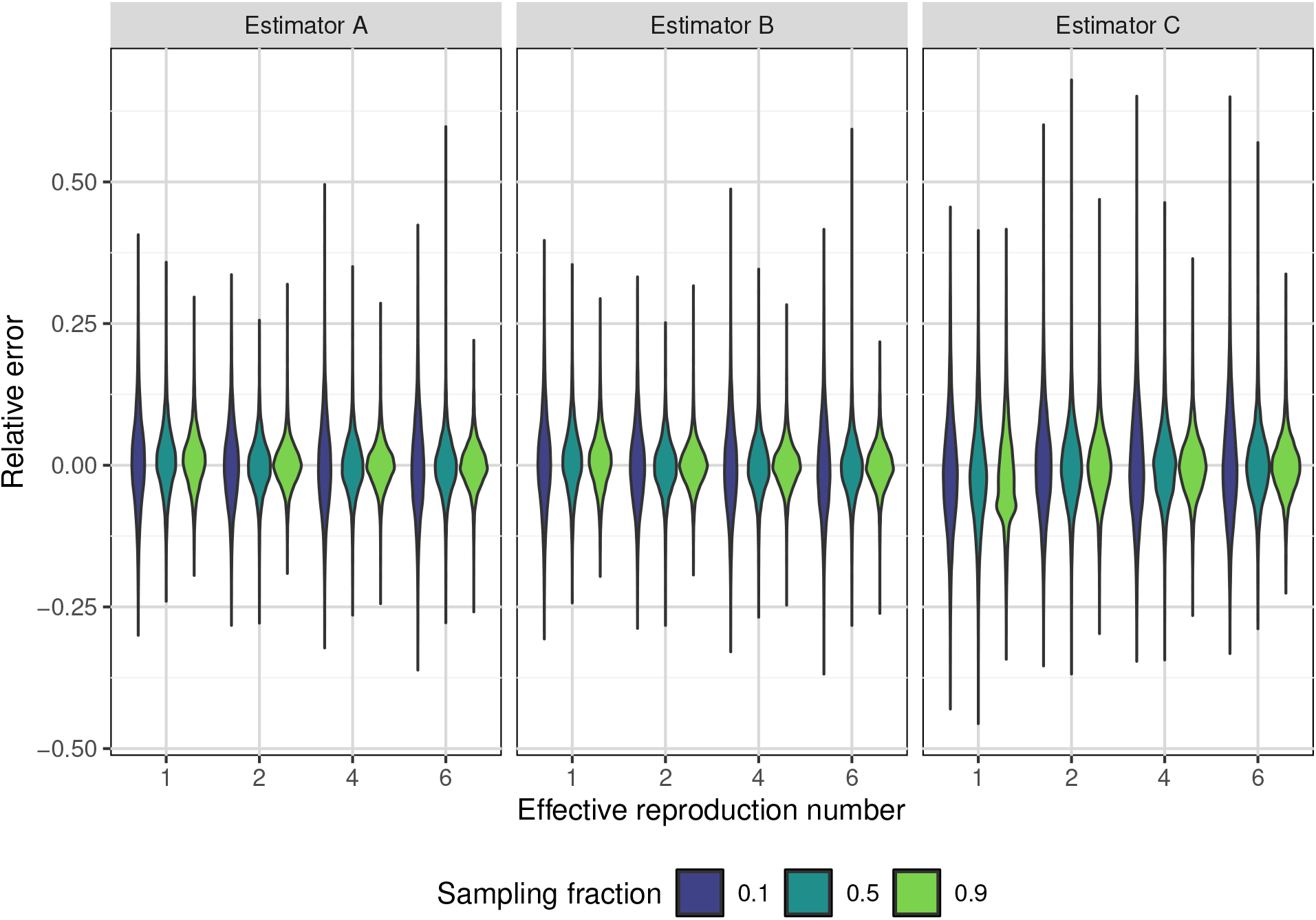
Relative error of the ECHO estimators under different combinations of *R*_*e*_ and *ρ*.

## S2 Justification of Estimator B

In ECHO Estimator B we note that each MIT individual will contribute a portion of their infectious period to the phylogeny, *τ*_drop_ *< τ*_*I*_ . However, the scale parameter used by Estimator B, 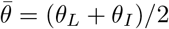, implicitly assumes that all MIT individuals contribute their full infectious interval on average. In this supplement we demonstrate why this works by deriving the conditional distribution of *τ*_*I*_ and *n*_off_, the number of secondary cases, given that an individual is MIT and show that the expected value of both is larger than that of their marginal distributions. If MIT individuals have a longer infectious interval on average, then the truncation of their infectious interval may be offset.

### Variables

- *n*_off_ – number of offspring/secondary cases
- *τ*_*I*_ – duration of the infectious period
- *n*_*s*_ – number of offspring that are sampled
- *ρ* – fixed sampling probability
- *r*_*e*_ – average number of secondary cases generated per day
- *θ*_*I*_ – average duration of the infectious period

### Distributions

- *τ*_*I*_ ∼ Exponential(*θ*_*I*_ )
- *n*_off_ | *τ*_*I*_ ∼ Poisson(*r*_*e*_*τ*_*I*_ )
- *n*_*s*_ | *n*_off_ ∼ Binomial(*n*_off_, *ρ*)

### S2.1 Average infectious intervals of MIT cases

We first show that MIT individuals will have longer infectious intervals on average.

#### S2.1.1 Conditional probability of a case being MIT given *τ*_*I*_

A case is MIT if they are unsampled and any of their descendant lineages are sampled. As this supplementary is intended to be a sketch, we simplify this to just their direct offspring being sampled. In general, this definition would produce a kernel similar to that provided in [8], but is beyond the scope of this supplement. Additionally, in the absence of extreme sampling heterogeneity, the sampling fraction should be roughly constant for the secondary cases of a particular focal case.

To derive *P*(MIT | *τ*_*I*_ ), the probability that a case is MIT given the duration of their infectious interval, we sum over the distribution of *n*_off_:

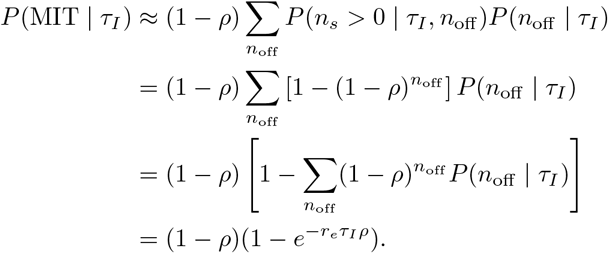

Therefore, the conditional distribution is

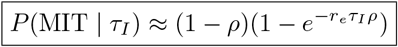

This confirms the initial intuition: if *τ*_*I*_ is larger, the probability of being MIT is larger. And similarly if *ρ* is large.

#### S2.1.2 Marginal distribution of being MIT

Given *P*(*MIT* |*τ*_*I*_ ), we can then derive the marginal probability of a case being MIT.

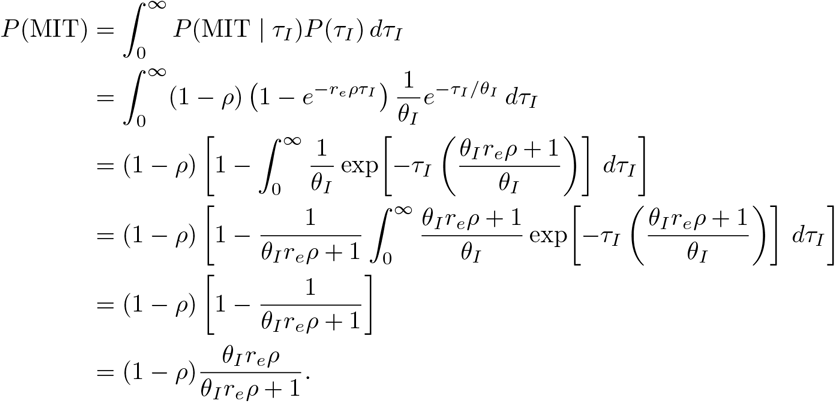

Substituting *R*_*e*_ = *θ*_*I*_ *r*_*e*_, gives

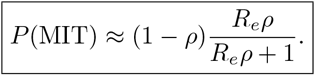

So, if *R*_*e*_ increases, then the probability that a case is MIT increases (and therefore so does the total number of MIT cases). The opposite is true if *ρ* increases. This is also consistent with intuition.

#### S2.1.3 Conditional distribution of *τ*_*I*_ given that a case is MIT

To determine whether or not MIT cases have longer infectious intervals on average, we derive *P*(*τ*_*I*_ | MIT), the corresponding probability density.

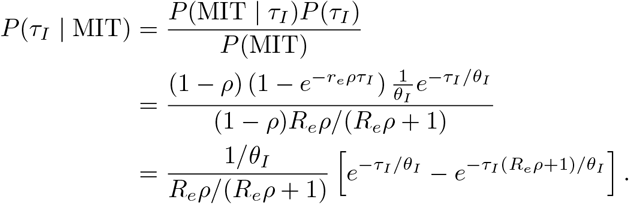

This density function is equivalent to a hypoexponential distribution (the sum of two exponential distributions) with means *θ*_*I*_ and 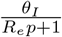. Therefore, the conditional average of *τ*_*I*_ for MIT cases is

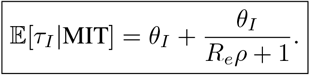

So, on average MIT individuals have longer infectious periods, on average multiplied by a factor of 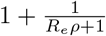.

### S2.2 Average number of secondary cases from MIT case

Since an MIT case is MIT if their descendant lineages are sampled, one should expect that focal cases with more secondary cases are more likely to be MIT. Therefore, we show in this section that conditional on an individual being MIT, their secondary case distribution has a higher average than the marginal secondary case distribution. The combination of the number of secondary cases and the sampling fraction should be related to *τ*_drop_, so it is important to understand how the distribution of secondary cases changes when we condition on being MIT.

#### S2.2.1 Conditional distribution of *n*_off_ given a case is MIT

First, note that the probability of a case being MIT, given the number of secondary cases, is 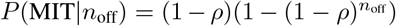. To obtain the marginal secondary case count distribution, integrate the joint distribution of *n*_off_ and *τ*_*I*_ with respect to *τ*_*I*_ :

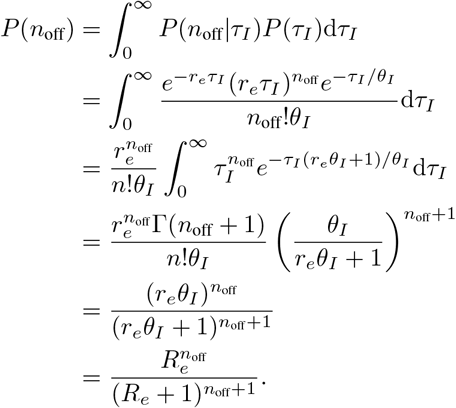

So, the marginal secondary case count distribution is a geometric distribution with probability *p*^*^, where 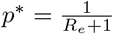.

Taking the distributions *P*(*n*_off_), *P*(MIT *n*_off_), and *P*(MIT), we can then write down the probability mass function for the conditional secondary case distribution of MIT individuals:

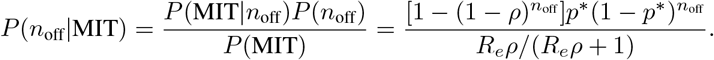

This does not correspond to a standard family of distributions, so the mass functions are illustrated in Figure S9. For this illustration we fix *R*_*e*_ = 2 and vary *ρ* = 0.1, 0.5, 0.9. The conditional distributions place more weight on larger values of *n*_off_ than the marginal and this tendency increase as *ρ* decreases. Correspondingly, E[*n*_off_ MIT] are 4.5 (*ρ* = 0.1), 3.5 (*ρ* = 0.5), and 3.07 (*ρ* = 0.9).

**Figure S9:**
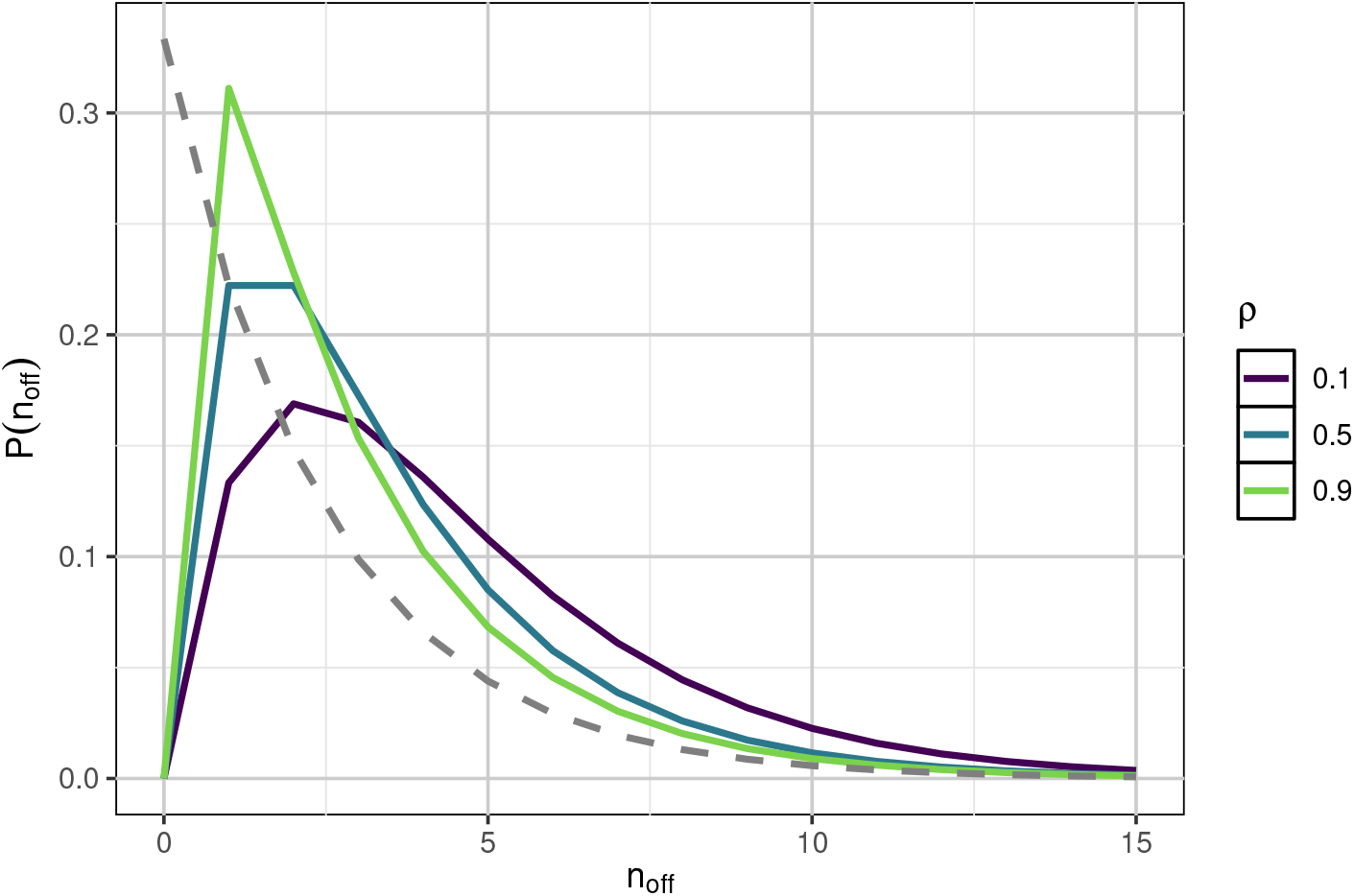
Solid lines are the probability mass function of *n*_off_ | MIT under different sample fractions (*ρ*). The grey dashed line is the marginal distribution of *n*_off_.

### S2.3 An empirical validation of Estimator B

In the preceding two sections we showed that both the duration of the infectious interval and the secondary case counts are more likely to be larger when an individual is MIT. However, given that these conditional distributions, and their expectations, depend on both *R*_*e*_ and *ρ*, we cannot fully justify the form of 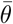 used by Estimator B. To complement the above derivation, we provide an additional experiment providing empirical support. In general, since *τ*_drop_ *< θ*_*I*_, we could formulate the likelihood of Estimator B as

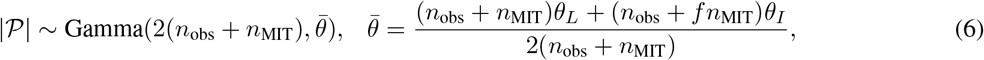

where *f* is some fraction chosen such that the average portion of an MIT individuals infectiousness is *fθ*_*I*_ . Figure S10 below shows errors generated by ECHO Estimator B using three values of *f*: 1*/*2 (*τ*_drop_ occurs halfway through *τ*_*I*_ ), 1 (what ECHO uses), and *R*_*e*_*/*(*R*_*e*_ + 1) (approximately the average time of the last secondary infection). These experiments are run with *θ*_*L*_ = 10 and *θ*_*I*_ = 8, while varying *R*_*e*_ (values of 1, 2, 4, and 6) and the sampling fraction (values of 0.1, 0.5, 0.9). The results from using *f* = 1 are accurate on average, regardless of the value of *R*_*e*_ or the sampling fraction.

**Figure S10:**
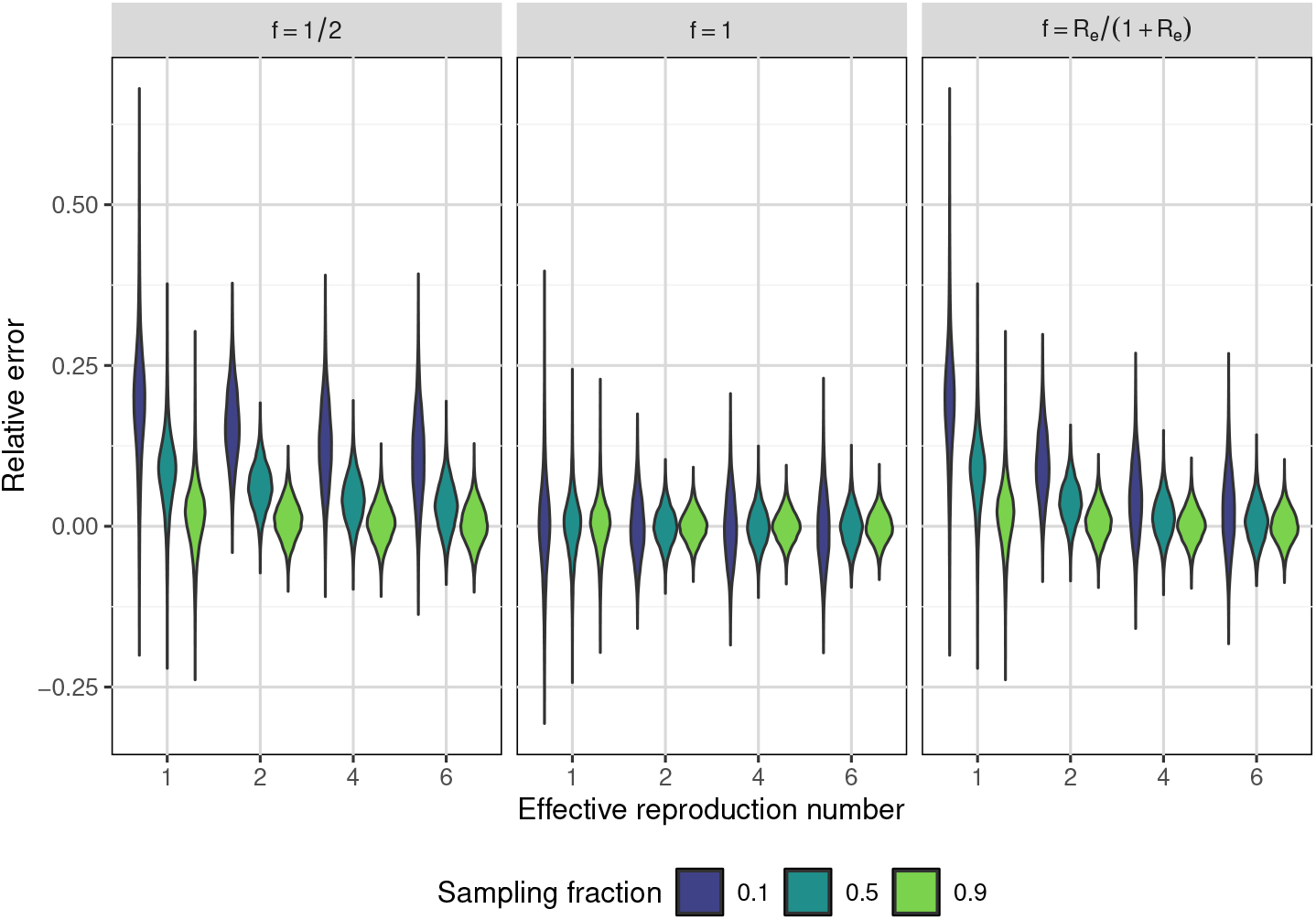
Relative error of ECHO Estimator B when using Equation 6. The experiments were run using *θ*_*L*_ = 10, *θ*_*I*_ = 8. Values for *R*_*e*_ and the sampling fraction are shown in the figure.

## Acknowledgments

We have received funding from the Wellcome Trust through the ARTIC2.0 network (award 313694/Z/24/Z), from the Canadian Natural Sciences and Engineering Research Council (NSERC RGPIN-2025-04335) and from the Canada Research Chair program (CRC-2023-00335). We also thank the authors of Masters et al [12] for publicly sharing the measles data.

